# Impact of amoxicillin shortage on pneumococcal resistance and IPD in children: evaluation of different management strategies in European countries

**DOI:** 10.1101/2025.04.15.25325803

**Authors:** Aurélie Maurin, Tristan Delory, Josselin LE Bel, Didier Guillemot, Mircea T. Sofonea, Laura Temime, Lulla Opatowski

## Abstract

Antibiotic shortages are increasing worldwide, with potential major consequences for both individual health and bacterial ecology. Here, we assess the impact of beta-lactam shortage management on pneumococcal resistance and the incidence of invasive pneumococcal disease (IPD).

We developed a mechanistic model of *S. pneumoniae* paediatric colonisation and transmission, accounting for beta-lactam and macrolide exposure. We explored the effects of four antibiotic allocation strategies following a one-year beta-lactam shortage: lowering consumption frequency, shortening treatment duration, reducing the daily dose, or substituting beta-lactams with macrolides. These strategies were analyzed in different European pharmaco-epidemiological settings.

Our findings reveal heterogenous impacts of allocation strategies, amplified at high shortage intensity. Shortage-induced consequences increased with baseline antibiotic consumption levels. Reducing beta-lactam consumption frequency was the most effective approach to managing pneumococcal resistance across Europe, decreasing penicillin-non-susceptible and multidrug-resistant strains by up to -21.4% in Spain, for a 50% shortage. The optimal strategy for minimizing IPD incidence was country-dependent: either lowering the daily dose or beta-lactam-to-macrolide substitution. However, the latter significantly increased macrolide resistance, with a relative rise by up to 26.2% in Denmark, for a 50% shortage.

Our results show that public health priorities and country-specific pharmaco-epidemiological factors should guide antibiotic management strategies during antimicrobial shortages.

## INTRODUCTION

*Streptococcus pneumoniae* (pneumococcus), a common human commensal bacteria, is the leading cause of morbidity and mortality among children under five years of age^1^. The most severe forms of disease, known as invasive pneumococcal disease (IPD), causes over 300,000 deaths annually in children under five worldwide^2^. In addition, antimicrobial resistance in pneumococcus is a major global health concern, as more than 20% of pneumococcal paediatric isolates^3^ worldwide are non-susceptible to first-line antibiotics, including amoxicillin. In this context, macrolide-resistant *S. pneumoniae* has been identified by the WHO as a bacterial priority pathogen^2^.

Alongside resistance, many countries face recent issues of antibiotic shortages, with the most severe deficits observed in generic medications^4^. For instance, the number of antibiotic shortages reported to the French National Drug Safety Agency (ANSM) rose from 64 in 2018 to 153 in 2020^5^, with beta-lactam antibiotics accounting for 52% of all reported or at-risk shortages^5^. Since October 2022, severe supply shortages of amoxicillin paediatric forms either alone or in combination with clavulanic acid, have been reported in Europe, the US and Asia, ^6–9^. Amoxicillin, a first-line therapy for most paediatric bacterial invasive infections^10^, is an effective and relatively inexpensive beta-lactam drug available in generic formulations. Given its widespread need and use in treating various paediatric diseases, the shortage of amoxicillin is a key public health issue with potentially life-threatening consequences.

Amoxicillin shortages have direct effects, hindering timely access to effective therapies, and indirect effects, influencing prescription practices by encouraging shorter treatment durations and promoting more rational antibiotic consumption and use of second-line agents^7,8,11,12^. For instance, studies have documented the impact of the 2022-to-2023 amoxicillin shortage on prescription practices, revealing a decrease of approximately 70% in amoxicillin prescription among institutions from the Sharing Antimicrobial Reports for Pediatric Stewardship OutPatient ^7^ and a 91% prescription reduction for children diagnosed with acute otitis media in a single-center study in the US^8^. As a consequence, antibiotic shortages may increase the risk of diseases^13^ but also constitute a threat in terms of emergence and spread of antimicrobial resistance^14^. Notably, given that antibiotic consumption is one of main drivers of resistance patterns in *S. pneumoniae*^15^, amoxicillin shortages raise concerns regarding pneumococcal resistance and carriage prevalence^8,14^.

However, the exact impact of antibiotic shortages on pneumococcal resistance is unknown, and the extent to which antibiotic allocation patterns in the context of a shortage affect pneumococcus ecology has not been studied. Here, we used dynamic mathematical modelling to investigate this interplay and the impact of beta-lactam shortage on pneumococcal resistance and IPD through a range of antibiotic use strategies in response to antibiotic shortage. By analysing different prescribing patterns and their effects on resistance and disease incidence, we seek to provide insight that can inform clinical guidelines and public health policies to better manage future antibiotic shortages and their consequences.

## RESULTS

We developed a deterministic susceptible–colonized–susceptible model of *S. pneumoniae* transmission in children under five years of age, considering possible exposure to two distinct antibiotic classes: beta-lactams and macrolides (**Fig. 1**). Children are described according to their antibiotic exposure and colonisation status. The model includes 16 distinct statues of *S. pneumoniae* colonisation, based on strain’s penicillin resistance levels (characterized by their minimum inhibitory concentrations (MIC) ranging from 0.06 to 8 mg/Liter), and macrolide resistance (susceptible or resistant). We assume that resistant strains incur a fitness cost, making them less transmissible. Individuals are exposed to antibiotics independently of their colonisation status. The model incorporates drug dosing effects by specifying a decolonization rate based on the antibiotic doses administered and the strain’s resistance level. Exposed individuals cannot acquire the sensitive strain of the corresponding antibiotic class in the model. Colonized children may develop invasive pneumococcal infections (IPDs) at rate of 4.8 × 10^−7^per day during colonisation.

**Fig. 1.**
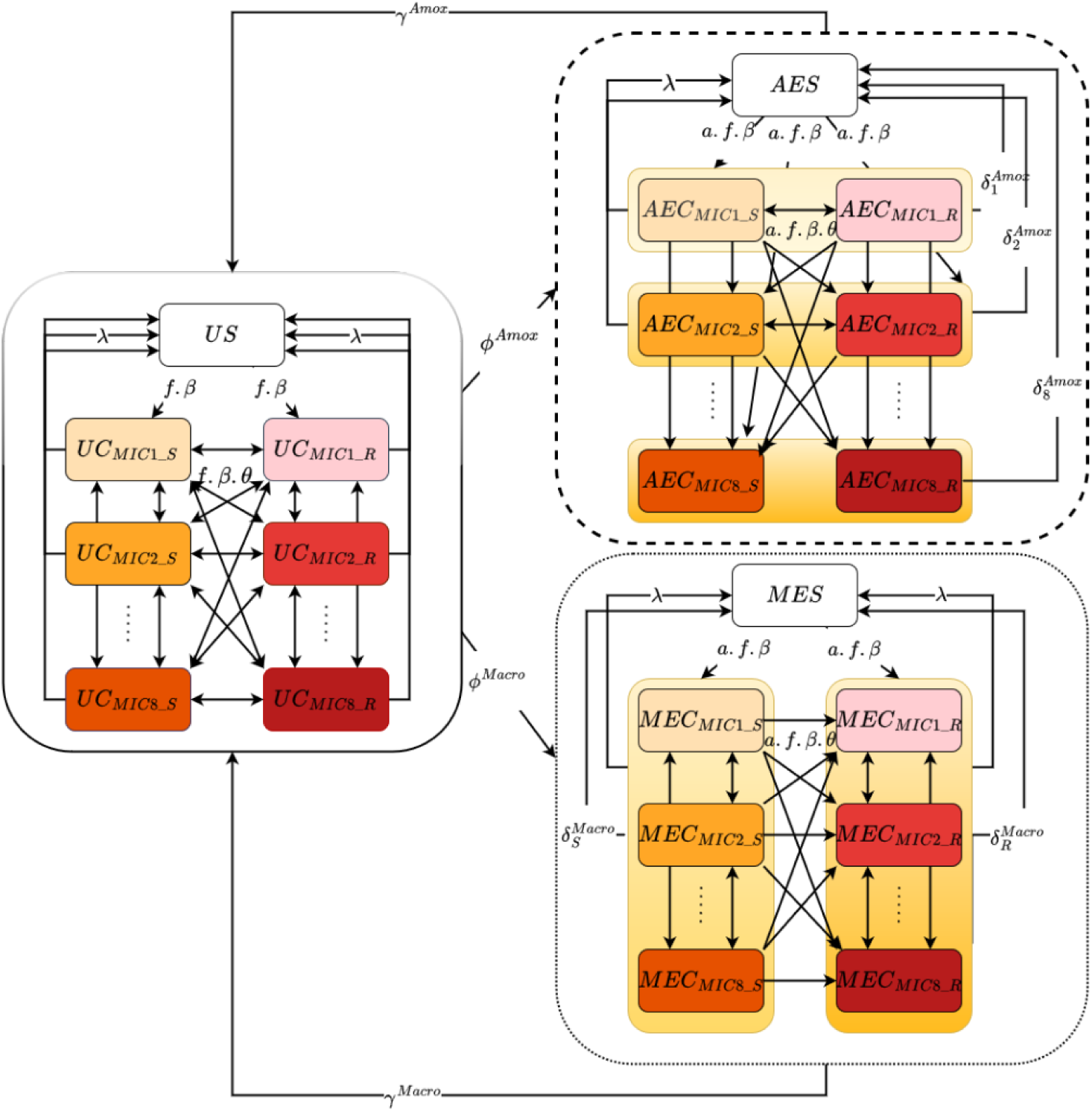
Transmission model of pneumococcus in children under five years old. The children population was divided according to antibiotic exposure: unexposed (U), beta-lactam exposed (AE), macrolide exposed (ME); pneumococcal colonization status : non-carrier (S) and carrier (C); and, among carriers, different levels of resistance to both beta lactams, according to the minimum inhibitory concentration (MIC), and macrolides, described as Susceptible or Resistant : *C*_*CMIk*_*m*_, *k* ϵ {*CMI*1, *CMI*2, …, *CMI*8} *and m* ϵ {*S*, *R*}. Children can be exposed to beta-lactams at rate ϕ^*Amox*^or to macrolides at rate ϕ^*Macro*^. Antibiotic exposure ends at a rate γ^*Amox*^and γ^*Macro*^, respectively. β represents the pneumococcal transmission rate, multiplied by a fitness cost *f* depending on the level of strain resistance. Replacement in colonization is possible but is penalized in transmission with a competition parameter θ. In children exposed to antibiotics, we assume that acquisition of strains susceptible to the administered antibiotic is impossible.

In the rest of this article, “penicillin-susceptible *S. pneumoniae”* refers to strains with an MIC ≤ 0.063 mg/L, “penicillin-intermediate” to those with 0.063 <MIC ≤2 mg/L, and “penicillin-resistant” to those with MIC > 2 mg/L. The term “penicillin-non-susceptible *S. pneumoniae”* includes both intermediate and resistant strains (I+R). “Macrolide-resistant *S. pneumoniae*” refers to strains with an inhibition zone < 19mm when testing with erythromycin or clindamycin.

Throughout this article, “beta-lactam antibiotics” (ATC: J01C) refer to penicillins and other beta-lactam antibacterials. As amoxicillin and amoxicillin-clavulanate account for more than 90% of all beta-lactam antibiotic sales in the European pharmaceutical market^16^, we will refer to amoxicillin shortage as beta-lactam shortage in the rest of the article. “Macrolide antibiotics” (ATC: J01F) refer to macrolides, lincosamides and streptogramins.

### The strategies adopted following antibiotic shortage lead to different population patterns

We explored four different responses to antibiotic shortages: reducing beta-lactam consumption frequency (strategy S1), reducing beta-lactam treatment duration (strategy S2), maintaining the frequency of beta-lactam consumption while reducing the daily dose (strategy S3) and switching from beta-lactam to macrolides prescription (strategy S4) in the French context. Note that the observed effects of strategy S1 could be due either to physicians deliberately reducing prescriptions in response to the shortage, or to patients struggling to access their prescribed treatments due to lack of availability. These strategies, representing the same global quantity of antibiotic distributed in the paediatric population, have different effects on the percentage of children exposed at each time and on prescribed doses (**Fig. *2*a**). The DDD values used to illustrate the impact of the different strategies on dose are 1.5 g for amoxicillin in the beta-lactam class and 2 g for pristinamycin in the macrolide class, based on the WHO standards^17^. Strategy S1 decreases the incidence and prevalence of beta-lactam-exposed children, as well as the number of amoxicillin DDD per 1,000 children and per day. Strategy S2 leads to a decrease in both the prevalence of beta-lactam-exposed children and the number of amoxicillin DDD per 1,000 children and per day, with no change in exposure incidence. Strategy S3 results in a decrease of the number of amoxicillin defined daily doses (DDD) per 1,000 children and per day, without affecting the incidence and prevalence of beta-lactam-exposed children. These first three strategies have no impact on macrolide exposure. Conversely, strategy S4 leads to reductions in all beta-lactam-related outcomes and to an increase in the incidence and prevalence of macrolide-exposed children, as well as in the number of pristinamycin DDD per 1,000 children and per day.

**Fig. 2.**
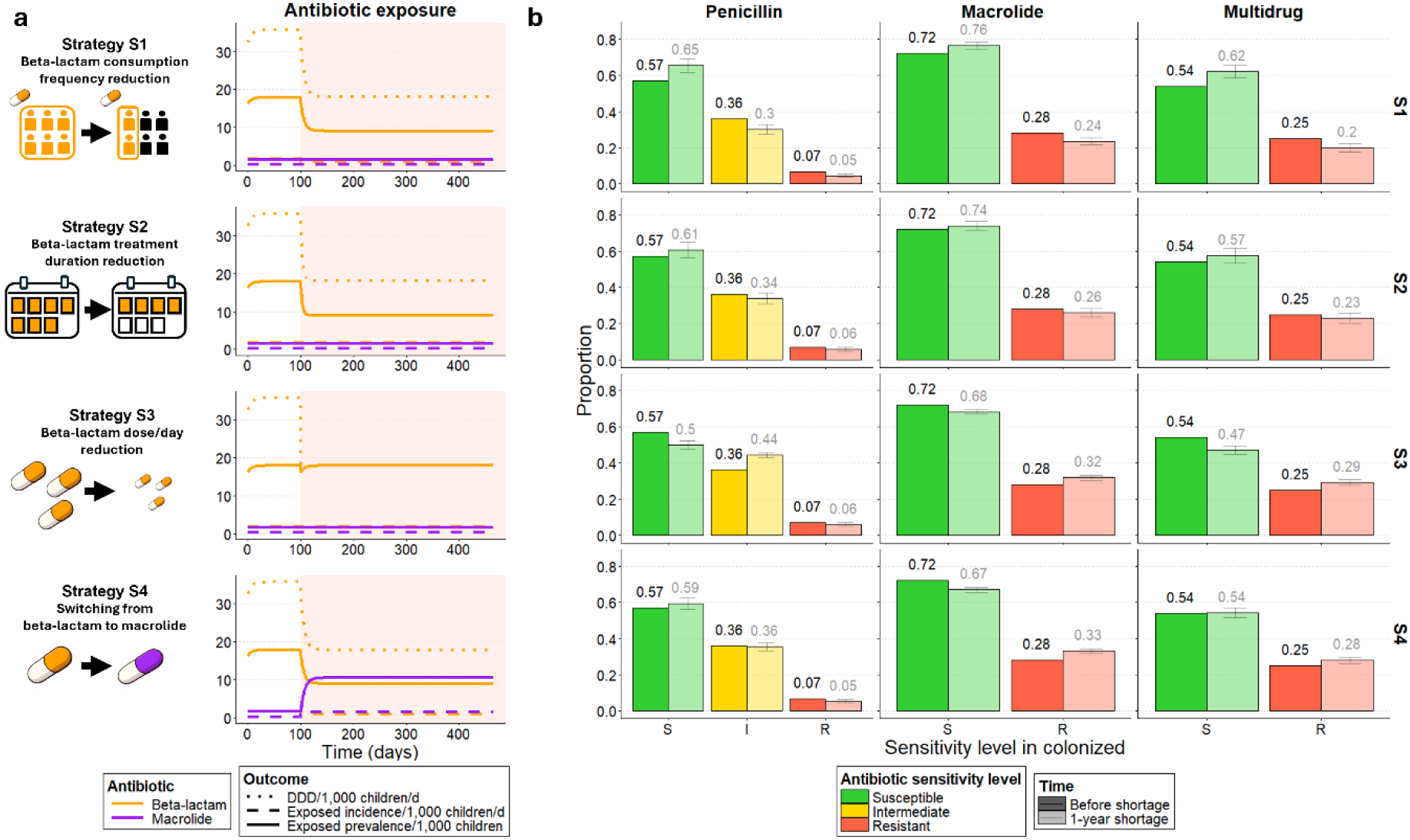
Impact of different shortage management strategies on the evolution of antibiotic exposure and pneumococcal resistance, assuming a 1-year long 50% beta-lactam shortage in the French context. 4 strategies are explored: reducing beta-lactam consumption frequency (S1), reducing beta-lactam treatment duration (S2), reducing beta-lactam daily dose (S3), switching from beta-lactam to macrolide prescription (S4). a) Impact on the defined daily dose (DDD) per 1,000 children per day, the daily incidence in antibiotic initiation of treatment and the prevalence of exposed children to beta-lactams and macrolides, the two considered antibiotic classes. The DDD values used are 1.5 g for amoxicillin and 2 g for pristinamycin, based on the WHO standards^17^. The colored area corresponds to the 1-year shortage period. b) Predicted proportion of penicillin-susceptible, penicillin-intermediate, penicillin-resistant, macrolide-susceptible, macrolide-resistant and multidrug-resistant *S. pneumoniae* strains in carriage at a specific time point, after one year of shortage, for the different strategies. The results are shown accounting for an uncertainty in the duration of carriage and the initial carriage prevalence. For each outcome and strategy, the barplot provides the median, with range bars representing the range from minimum to maximum values.

As a consequence of changes in antibiotic exposure, the evolution of bacterial ecology during a 1-year shortage also depends on the adopted strategy (**Fig. *2*b**). Strategies S1 and S2 lead to a relative decrease in the proportion of both penicillin-intermediate (−16.6% [-24.2%,-9.4%] and - 6.4% [-15.3%, 1.8%]; median [min, max]) and penicillin-resistant strains (−34.2% [-48.2%, -20.1%] and -18.1% [-35.3%, -1.3%]). S1 and S2 also reduce the proportion of macrolide-resistant *S. pneumoniae* (MRSP, -15.7% [-23.6%, -8.4%] and -6.7% [-15.7%, 1.7%]) and multidrug-resistant *S. pneumoniae* (MDRSP, -19.4% [-28.6%, -10.6%] and -8.3% [-19.1%, 1.9%]). Strategy S3 causes a relative decrease in the proportion of both penicillin-susceptible *S. pneumoniae* (PSSP, -12.7% [-16.7%, -8.6%]) and penicillin-resistant strains (−13.2% [-29.1%, 1.8%]), but a relative increase in the proportion of penicillin-intermediate strains (+22.7% [19.0%, 26.1%]). S3 also leads to a relative rise in MRSP proportion (+13.4% [8.2%, 18.5%]) and MDRSP proportion (+16.6% [10.5%, 22.8%]), due to the initial presence of multi-resistance in a large part of circulating strains, as is the case in all European countries in 2021^18^ (see Supplementary Figure 1 for results obtained when assuming no multi-resistance). Strategy S4 leads to a relative increase in PSSP proportion (+3.8% [-1.6%, 9.5%]), MRSP proportion (+18.1% [13.0%, 23.0%]), and MDRSP proportion (+12.6% [5.0%, 19.8%]), while reducing both penicillin-intermediate (−1.6% [-7.8%, 4.1%]) and penicillin-resistant strains (−22.9% [-37.7%, -8.6%]).

### Shortage intensity’s impact on resistance and severe disease burden

We studied how increasing beta-lactam shortage intensity, from 0% to 95%, impacted pneumococcal epidemiology compared to no shortage (**Fig. *3***). Four epidemiological outcomes were studied over 1 year: the proportions of penicillin non-susceptible *S. pneumoniae* (PNSP), macrolide-resistant *S. pneumoniae* (MRSP), and multidrug-resistant *S. pneumoniae* (MDRSP) among all *S. pneumoniae* carried by colonized children; and the overall IPD incidence in these children, simply estimated as a fixed proportion of pneumococcal carriers. The largest relative increases are observed for PNSP and MRSP proportions under the daily dose reduction strategy S4 while the largest relative decreases are obtained for PNSP, MRSP and MDRSP proportions under the consumption frequency reduction strategy S1.

**Fig. 3.**
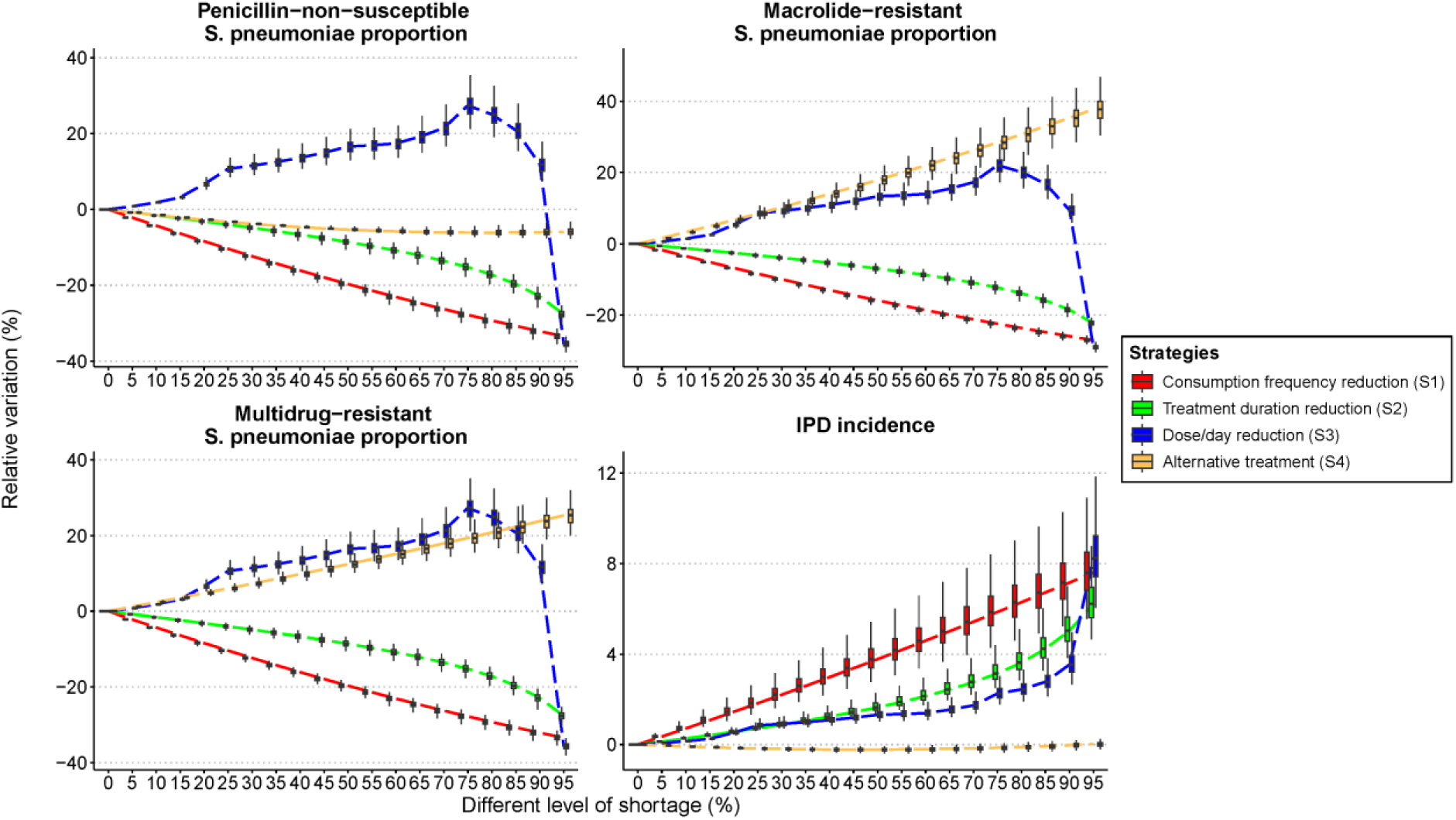
1-year impact of different shortage levels and management strategies on antibiotic resistance and the incidence of invasive pneumococcal diseases (IPD) in the French context. 4 strategies are explored: reducing beta-lactam consumption frequency (S1, red), reducing beta-lactam treatment duration (S2, green), reducing beta-lactam daily dose (S3, blue), switching from beta-lactam to macrolide prescription (S4, yellow). Shortages levels from 0% to 95% (x-axis) are explored, representing the availability reductions of beta-lactam quantity. Four outcomes are provided: (a-c) relative variation in the proportion of antibiotic-resistant proportion compared with no shortage: (a) penicillin-non-susceptible *S. pneumoniae* (PNSP) proportion, (b) macrolide-resistant *S. pneumoniae* (MRSP) proportion, (c) multidrug-resistant *S. pneumoniae* (MDRSP) proportion; (d) relative variation of overall invasive pneumococcal disease (IPD) incidence per 100,000 children per year compared with no shortage. The results are shown accounting for an uncertainty in the duration of carriage and the initial carriage prevalence. For each outcome and strategy, boxplots provide the median (black horizontal line), the interquartile range (upper and lower bounds of the box), and minimum and maximum values (whiskers).

All four outcomes vary quasi-linearly with shortage intensity up to 75% of shortage regardless of the applied strategy. For instance, relative variations in PNSP proportion range from -10.4% [-11.-%, -9.5%] to +10.7% [+8.4%, 14.8%] for a 25% shortage, from -19.7% [-21.6%, -18.2%] to +16.5% [12.9%, 23.0%] for a 50% shortage, and from -27.9% [-29.9%, -26.0%] to +27.4% [21.2%, 38.5%] for a 75% shortage, compared to no shortage after 1-year. Beyond 75%, the daily dose reduction strategy S3 causes a sharp drop in resistance while triggering an exponential increase in IPD incidence, with the daily dose administrated no longer sufficient to eliminate susceptible strains. The treatment duration reduction strategy S2 also shows an accelerating impact, breaking the linear trend. In contrast, strategies S1 and S4 maintain a quasi-linear relationship between shortage intensity and the burden of resistance and IPD across all shortage levels.

### Recommended strategies depends on the targeted outcome

Assuming a 50% beta-lactam shortage over a year, we aimed at identifying the strategies that minimize the burden of seven outcomes related to resistance and invasive diseases (**Fig. *4***). Simulations over a year show that the consumption frequency reduction strategy S1 is optimal for minimizing antibiotic resistance in the investigated ranges of carriage duration and initial prevalence. S1 results in a relative decrease of -19.7% in PNSP proportion (from 43% to 35% of all carried pneumococci), -15.9% in MRSP proportion (from 28% to 24% of all carried pneumococci), and -19.7% in MDRSP proportion (from 25% to 20% of all carried pneumococci), compared with no shortage. This strategy also minimizes the incidence of associated resistant *S. pneumoniae* IPDs, with a relative decrease of -6.9% in penicillin-non-susceptible IPD (*IPD*_*PNSP*_) incidence (from 3.7 to 3.4 infections per 100,000 children per year), -4.8% in macrolide-resistant IPD (*IPD*_*MRSP*_) incidence (from 2.4 to 2.2 infections per 100,000 children per year), and -6.9% in multidrug-resistant IPD (*IPD*_*MDRSP*_) incidence (from 2.1 to 1.9 infections per 100,000 children per year) (**Fig. *4***). The duration reduction strategy S2 also leads to reductions, but to a lesser extent than S1 (between 2.5 and 2.8-fold weaker reductions).

**Fig. 4.**
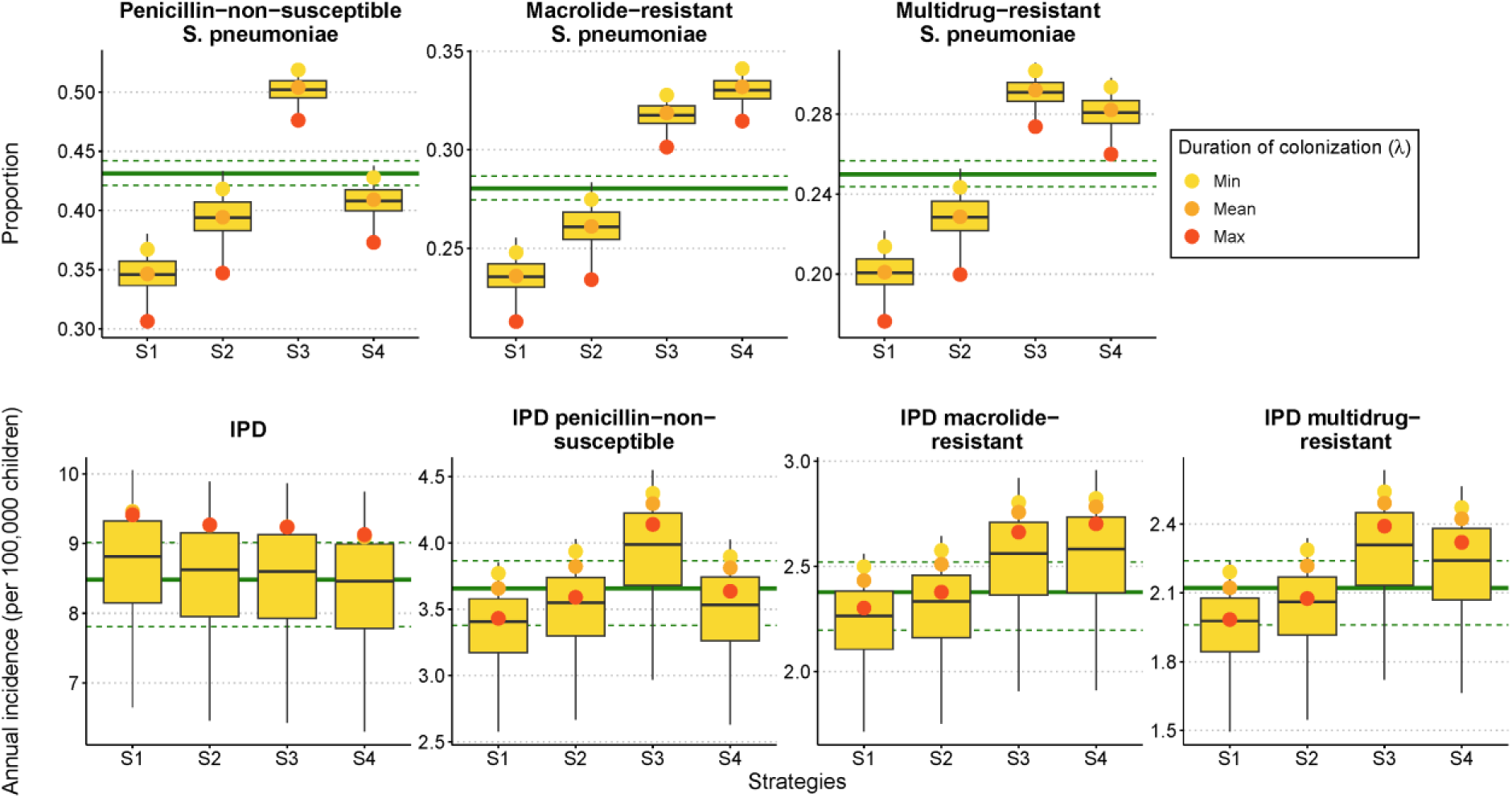
1-year impact of a 50% beta-lactam shortage on antibiotic resistance and invasive pneumococcal disease (IPD) incidence, depending on antibiotic shortage management strategies in the French context. Assuming a 50% shortage, 4 strategies are explored: reducing beta-lactam consumption frequency (S1), reducing beta-lactam treatment duration (S2), reducing beta-lactam daily dose (S3), switching from beta-lactam to macrolide prescription (S4). Seven outcomes are provided: (a-c) relative variation in the proportion of antibiotic-resistant strains compared with no shortage : (a) penicillin-non-susceptible *S. pneumoniae* (PNSP) proportion, (b) macrolide-resistant *S. pneumoniae* (MRSP) proportion, (c) multidrug-resistant *S. pneumoniae* (MDRSP) proportion; (d-g) relative variation of invasive pneumococcal disease (IPD) incidence per 100,000 children per year compared with no shortage, (d) overall incidence (IPD), (e) penicillin-non-susceptible IPD incidence (*IPD*_*PNSP*_), (f) macrolide-resistant IPD incidence (*IPD*_*MRSP*_), and (g) multidrug-resistant IPD incidence (*IPD*_*MDRSP*_). The results are shown accounting for an uncertainty in the duration of carriage and the initial carriage prevalence. For each outcome and strategy, boxplots provide the median (black horizontal line), the interquartile range (upper and lower bounds of the box), and minimum and maximum values (whiskers). For each outcome, the bold green line depicts the median value obtained in the baseline scenario without beta-lactam shortage, and the dotted lines the associated interquartile range. Additionally, the yellow, orange and red dots correspond to the outcome values for the minimum (λ*=* 1/51 day^-1^), average (λ*=* 1/43 day^-1^) and maximum (λ*=* 1/32 day^-1^) duration of colonization values, with the carriage prevalence fixed at 52% of individuals.

To minimize overall IPD incidence, no single strategy is clearly better than the others. The antibiotic switching strategy S4 slightly limits this incidence, with a relative reduction of -0.21%. However, this strategy also induces a +17.9% increase in MRSP proportion (from 28% to 33% of all carried pneumococci) and a +12.4% increase in MDRSP proportion (from 25% to 28% of all carried pneumococci). Interestingly, the daily dose reduction strategy S3 is never advisable as it increases all indicators, resulting in a relative increase ranging from +1.3% in IPD incidence, and up to +16.5% in PNSP and MDRSP proportions (**Fig. *4***).

When considering invasive disease outcomes, the uncertainty range is broad and the advantage between strategies for the same outcome often overlaps. However, when the initial prevalence and duration of colonization are held constant, the advantage of one strategy over the others is consistent. Supplementary Figure 2 provides outcomes values across strategies for the maximum, mean and minimum values of the prevalence of carriage.

### Recommended strategies depend on pre-shortage epidemiology

In a partial rank correlation coefficients (PRCC) multivariate sensitivity analysis, we found that the key parameters to consider when selecting the optimal strategy in an beta-lactam shortage context were those describing the initial resistance epidemiology (*Rcr*^*init*^, *PNSP*^*init*^ and *MRSP*^*init*^), carriage prevalence (*pi*^*init*^), and treatment duration (γ^*Amox*^and γ^*Macro*^) (see Supplementary Figure 3 for the PRCC analysis). As these variables tend to differ in different settings, we explored how the initial epidemiology of resistance (*PNSP*^*init*^, *MRSP*^*init*^and *Rcr*^*init*^) influences the preference for one strategy over another, while holding the other parameters fixed.

*PNSP*^*init*^and *MRSP*^*init*^describe initial proportions of penicillin non-susceptible and macrolide-resistant strains, while *Rcr*^*init*^ corresponds to the initial proportion of macrolide-resistant strains among penicillin non-susceptible strains (i.e., multi-resistance). Variations in other parameters identified in the sensitivity analysis have less impact on the recommended strategies (see Supplementary Figure 4 for recommended strategy according to the value of *PNSP*^*init*^, *MRSP*^*init*^, *pi*^*init*^, λ, γ^*Amox*^and γ^*Macro*^). A complementary sensitivity analysis was conducted on the competition parameter θ, showing little impact on the outcome values, but an effect on the fitted fitness cost values (see Supplementary Figure 5 for the effect of the θ parameter).

When “minimising PNSP proportion” is the outcome of interest, the antibiotic switching strategy S4 comes out as best when multi-resistance is infrequent (*MRSP*^*init*^ ≥ *Rcr*^*init*^), while the consumption frequency reduction strategy S1 is selected in case of frequent multi-resistance (*MRSP*^*init*^ < *Rcr*^*init*^). The same conclusions apply for penicillin non-susceptible IPD (IPD_PNSP_), where either S1 or S4 is recommended based on *MRSP*^*init*^and *Rcr*^*init*^ values (**Fig. *5***). Considering the “minimization of MRSP proportion”, the daily dose reduction strategy S3 is recommended when multi-resistance is infrequent (*MRSP*^*init*^ ≥ *Rcr*^*init*^), whereas the strategy S1 is again advised in case of frequent multi-resistance (*MRSP*^*init*^ < *Rcr*^*init*^). For “macrolide-resistant IPD (IPD_MRSP_) minimization”, the recommended strategy varies among strategies S1, S2 and S3 depending on *MRSP*^*init*^, *Rcr*^*init*^and *PNSP*^*init*^values, though most differences are not significant.

**Fig. 5.**
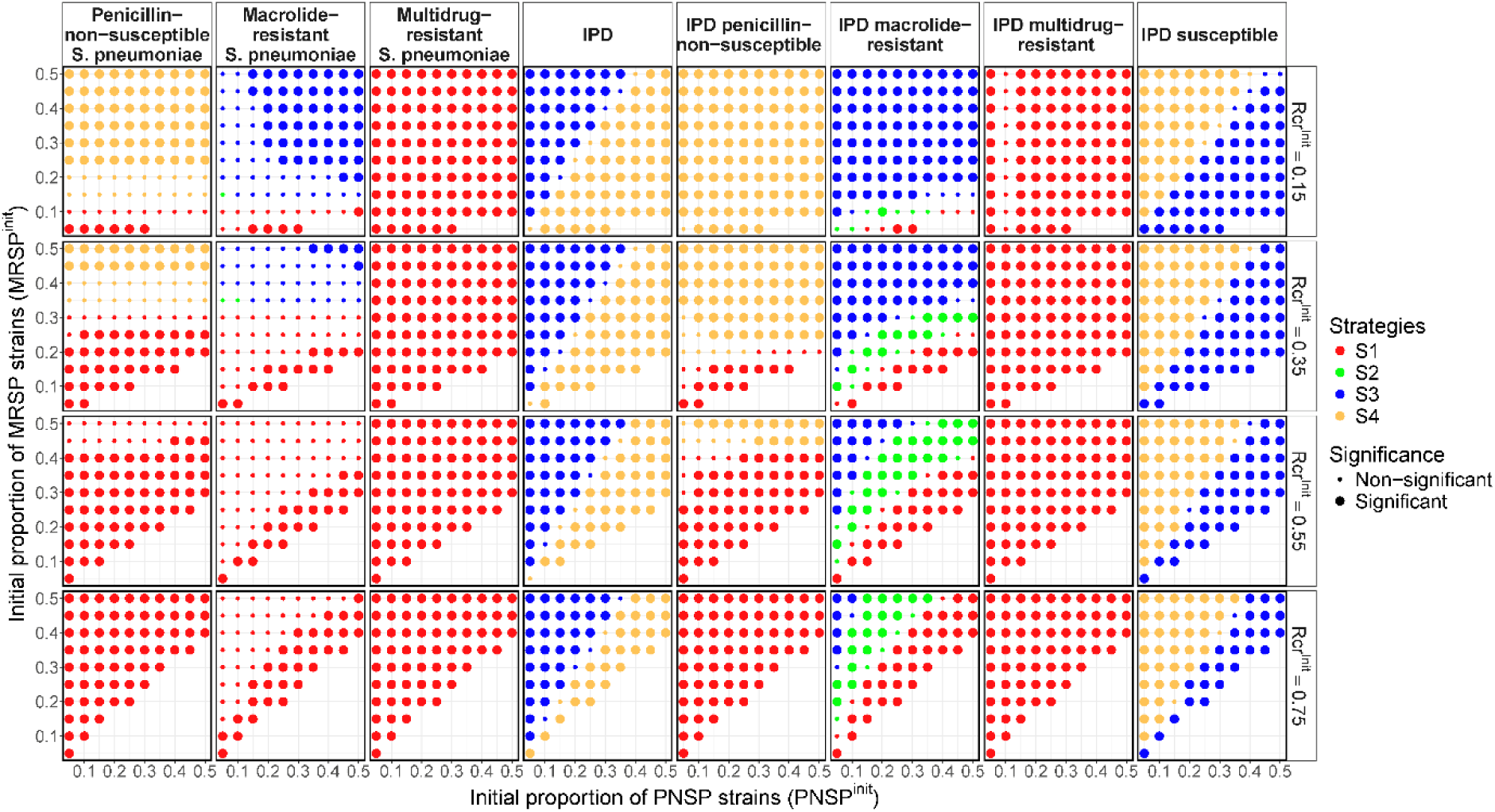
Recommended antibiotic shortage management strategy depending on the initial proportions of penicillin-non-susceptible strains (*PNSP*^*init*^), macrolide-resistant strains (*MR*^*init*^) and the proportion of macrolide-resistant strains among penicillin-non-susceptible strains (*Rcr*^*init*^) before shortage start, after a 1-year period of 50% beta-lactam shortage. 4 strategies are explored: reducing beta-lactam consumption frequency (S1, red), reducing beta-lactam treatment duration (S2, green), reducing beta-lactam daily dose (S3, blue), switching from beta-lactam to macrolide prescription (S4, yellow). Seven outcomes are provided: (a-c) relative variation in the proportion of antibiotic-resistant strains compared with no shortage : (a) penicillin-non-susceptible *S. pneumoniae* (PNSP) proportion, (b) macrolide-resistant *S. pneumoniae* (MRSP) proportion, (c) multidrug-resistant *S. pneumoniae* (MDRSP) proportion; (d-g) relative variation of invasive pneumococcal disease (IPD) incidence per 100,000 children per year compared with no shortage, (d) overall incidence (IPD), (e) penicillin-non-susceptible IPD incidence (*IPD*_*PNSP*_), (f) macrolide-resistant IPD incidence (*IPD*_*MRSP*_), (g) multidrug-resistant IPD incidence (*IPD*_*MDRSP*_) and (h) susceptible IPD incidence (*IPD*_*SSP*_). For each initial condition combination, the strategy minimizing each outcome is depicted with a coloured dot. A strategy is considered better (indicated by a large dot) when its value differs by more than 5% from the others for proportion-based outcomes, or by at least 1 IPD/1 million children/year for incidence-based outcomes.

S1 is consistently the recommended strategy when minimizing MDRSP or IPD_MDRSP_ is the outcome of interest. Finally, the best strategy to “minimize the overall IPD incidence” is the daily dose reduction strategy S3 when macrolide resistance is initially more frequent than penicillin non-susceptibility (*MRSP*^*init*^ > *PNSP*^*init*^), and the antibiotic switching strategy S4 otherwise (*MRSP*^*init*^ ≤ *PNSP*^*init*^) (**Fig. *5***). Conversely, to “minimize the incidence of susceptible IPD (IPD_SSP_)”, the best strategy is strategy S3 if *MRSP*^*init*^ ≤ *PNSP*^*init*^, and strategy S4 if *MRSP*^*init*^ > *PNSP*^*init*^.

### Application to shortages scenarios in European contexts

As an illustration, we finally applied the model to 20 European countries (Austria, Belgium, Croatia, Czechia, Denmark, Estonia, Finland, France, Germany, Hungary, Ireland, Italy, Lithuania, Netherlands, Norway, Poland, Portugal, Slovenia, Spain and Sweden), initialising the model with contexts reflecting their current levels of pneumococcal resistance and antibiotic consumption. After a 1-year period of 50% beta-lactam shortage, we find a strong correlation between national antibiotic consumption frequency and the magnitude of the impact of shortage on the different outcomes (see Supplementary Figure 6 for the correlation matrix). In Spain, one of the highest antibiotic consumers, the relative variation in resistance outcomes varies from - 21.4% to +24.3%, and the absolute variation in IPD outcomes ranges from -2 to +4 cases per million children per year, across all scenarios. In contrast, the Netherlands, the lowest consumer, exhibits relative resistance variations from -6.1% to +9.8%, and absolute IPD variations from -1 to +1 case per million children per year, reflecting more than a threefold difference between these countries (**Fig. *6***). For disease-related outcomes, low-antibiotic-consumption countries (e.g. Finland, Germany, the Netherlands, etc.) exhibit consistent trends, with strategies S3 and S4 showing no significant effect and strategies S1 and S2 causing a slight increase in sensitive infections, while high-antibiotic-consumption countries (e.g. France, Ireland, Italy, Spain, etc.) show varied responses to strategies.

**Fig. 6.**
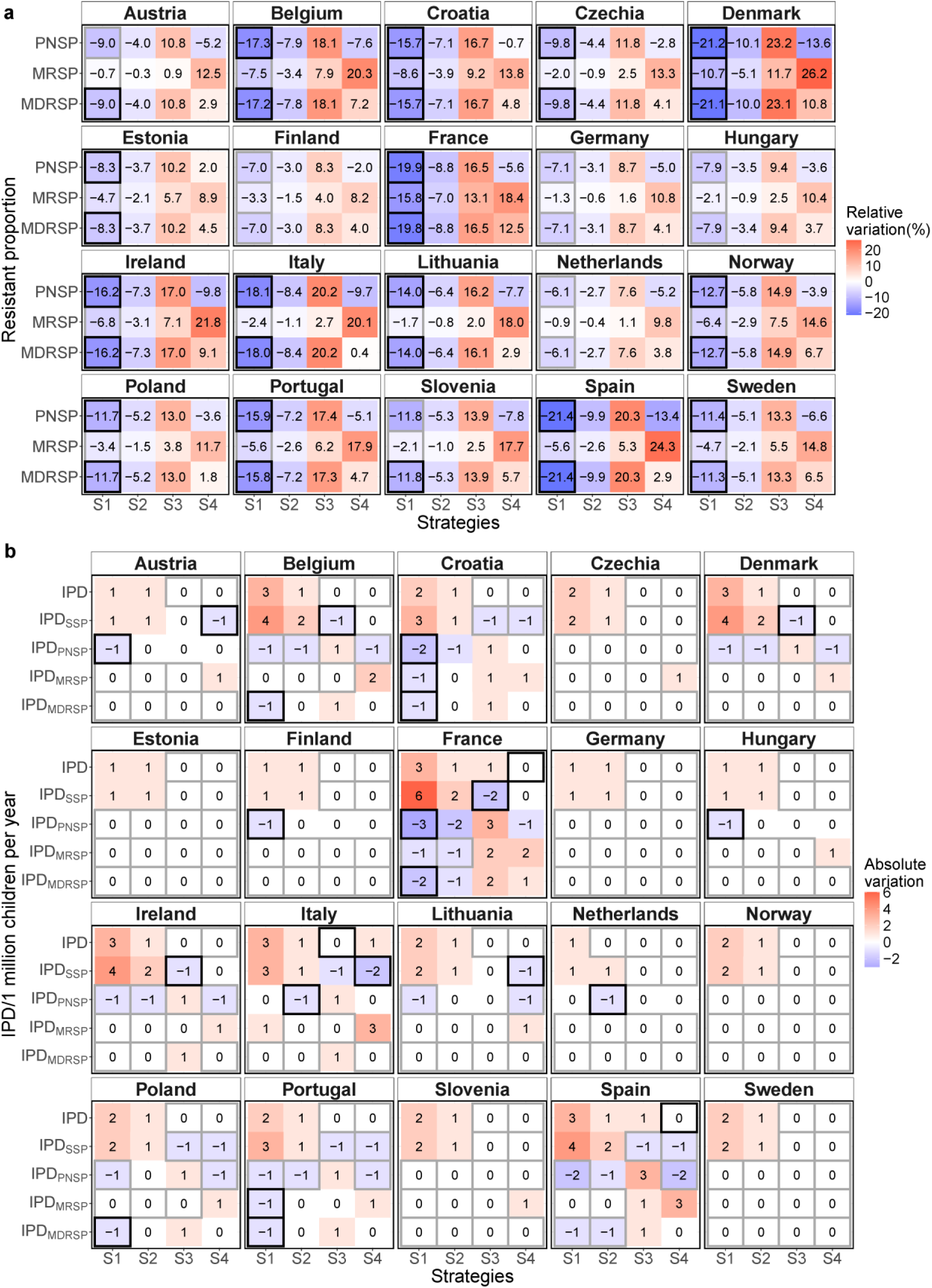
Variation in 20 European countries in *S. pneumoniae* resistance proportion and invasive disease incidence among children under five years of age, across different shortage management strategies following a 1-year period of 50% shortage compared to no-shortage. The model was informed with the 20 country-specific initial epidemiological conditions (initial proportion of penicillin-non-susceptibility, macrolide-resistance and multi-resistance) and antibiotic consumption frequency (baseline consumption of beta-lactams and macrolides) and run over 1 year. 4 strategies are explored: reducing beta-lactam consumption frequency (S1), reducing beta-lactam treatment duration (S2), reducing beta-lactam daily dose (S3), switching from beta-lactam to macrolide prescription (S4). The optimal strategy is framed. **a)** Three resistance proportion outcomes predicted: penicillin-non-susceptible *S. pneumoniae* (PNSP) proportion; macrolide-resistant *S. pneumoniae* (MRSP) proportion; and multidrug-resistant *S. pneumoniae* (MDRSP) proportion. For each outcome, black squares indicate the predicted optimal strategy for which the predicted proportion differs by more than 5% from all the other strategies, while grey lines indicate the best strategy, when the difference is lower. **b)** Five IPD outcomes are assessed/predicted/estimated: overall incidence (IPD);susceptible IPD incidence (*IPD*_*SSP*_); penicillin-non-susceptible IPD incidence (*IPD*_*PNSP*_) incidence; macrolide-resistant IPD incidence (*IPD*_*MRSP*_); and multidrug-resistant IPD incidence (*IPD*_*MDRSP*_). Black lines indicate the optimal strategy for which the predicted number of IPD cases differs from at least 1 case per million children per year from all the other strategies, while grey lines indicate the best strategy, but with a lower difference.

Strategy S1 (reducing beta-lactam consumption frequency) appears as the most effective strategy for managing antibiotic resistance across studied European countries. PNSP proportion decrease by a relative variation ranging from -6.1% in the Netherlands to -21.4% in Spain and the relative decrease in the MRSP proportion ranges from -0.9% in the Netherlands to -15.8% in France. However, across all countries, strategy S1 leads to an increase in overall IPD incidence (from 1 to 3 cases per million children per year), attributed to an increase in susceptible infections (from 1 to 6 IPD_SSP_ cases per million children per year) greater than the decrease in resistant infections (reduction of 1 to 3 IPD_PNSP_, 0 to 1 IPD_MRSP_, and 0 to 2 IPD_MDRSP_ cases per million children per year). Strategy S2 (reducing treatment duration) also increases the overall IPD incidence, with an increase in susceptible infections and a decrease in resistant ones, but its effects are about half as large as those of S1 across all countries. (**Fig. *6***). Supplementary Figure 7 depicts the absolute variations in disease outcomes for each specific resistance levels to both antibiotics.

While strategy S3 (reduction of beta-lactam daily dose) is not advisable for resistance-related outcomes, it limits overall IPD incidence (from 0 to 1 case per million children per year) and is substantially better than other strategies in countries where baseline macrolide-resistance exceeds penicillin non-susceptibility (*MRSP*^*init*^ > PNSP ^*init*^), such as Italy. S3 reduces the number of susceptible infections (reduction of 0 to 2 IPD_SSP_ cases per million children per year, across all countries) but leads to an increase in resistant infections (by 1 to 3 *IPD*_*PNSP*_, 0 to 2 *IPD*_*MRSP*_, and 0 to 2 *IPD*_*MDRSP*_ cases per million children per year, across all countries)(**Fig. *6***).

Finally, we forecast that switching from beta-lactam to macrolide prescription (strategy S4) results in a significant relative increase in the proportion of macrolide-resistant strains by a fraction ranging from +9.8% in the Netherlands to +26.4% in Denmark, after a 1-year 50% shortage. The impact of strategy S4 on MDRSP proportion is more varied, showing a +0.4% relative decrease in Italy and a +12.5% relative increase in France. S4 does not affect the overall IPD incidence and is substantially better than other strategies in countries where baseline macrolide-resistance is below penicillin non-susceptibility (where *MRSP*^*init*^ ≤ PNSP ^*init*^), such as France. Across all countries, S4 decreases IPD_SSP_ infections (by 0 to 2 cases per million children per year) but leads to an increase in resistant *IPD*_*MRSP*_ infections (from 0 to 3 cases per million children per year) (**Fig. *6***).

These results highlight that no single strategy fulfils all the criteria for minimizing both antibiotic resistance and severe infections incidence, making it essential to balance these outcomes when selecting the optimal strategy.

## DISCUSSION

Using a mathematical model of pneumococcal transmission that incorporates the effects of drug dosing on resistance levels for two antibiotic classes, we evaluated the impact of beta-lactam shortages under four alternative strategies. Our results showed that the impact on pneumococcal ecology varied widely depending on the chosen strategy, with stronger effects observed as the severity of the shortage increased. In particular, no single strategy consistently reduced both resistance and invasive disease, highlighting the need for tailored approaches that align with national public health priorities. Simulations across 20 European countries further showed that the effectiveness of the strategies was shaped by the initial pharmaco-epidemiological conditions, with more pronounced effects in countries with higher baseline antibiotic consumption frequency. Additionally, the optimal strategy for each outcome varied by country. This underscores the importance of considering country-specific factors when developing antibiotic stewardship policies.

Reducing beta-lactam daily dose was found to increase intermediate-resistant proportion at the population level due to the time-dependent action of the antibiotic. Indeed, lower defined daily doses have a lower efficacity against intermediate-resistant strains, allowing them to persist, as they are favoured over sensitive strains (which are easily eliminated) and resistant strains (which have a higher fitness cost). These findings are consistent with existing theoretical and observational studies showing that lower defined daily doses favour intermediate-resistant strains^19,20^. Simulations also showed that reducing the duration or the frequency of beta-lactam treatments in individuals reduce the proportion of PNSP while increasing the proportion of PSSP at the population level. In fact, both scenarios lead to a diminution in the instantaneous average number of individuals exposed to antibiotics, reducing selective pressure and favouring a return of susceptibility. This aligns with observational studies reporting reductions in the proportion of resistant strains following a decreased in antibiotic use in populations^21–23^. The antibiotic switching strategy (S4) affects the proportion of resistant strains through two synchronous mechanisms: a decrease in beta-lactam consumption and an increase in macrolide consumption. In our main analysis, because the prevalence of multi-resistance was high (20%) among circulating strains, these mechanisms overlap, resulting in stability in the proportion of penicillin-resistance (PNSP) and a significant rise in the proportion of macrolide-resistance (MRSP). Strong correlation between macrolide use and macrolide resistance was previously reported in a Finnish study^24^. In France, the 23% increase in macrolide use over 2019-2020^25,26^ was followed by a 15% increase in erythromycin-resistant strains^16^.

We highlight here that the expected impact of beta-lactam shortages on both resistance and incidence outcomes depends on country’s specific resistance and exposure characteristics. This is consistent with studies showing the importance of using local resistance data in the development of effective antibiotic stewardship strategies^28^. Regarding IPD incidence, we found that the effectiveness of switching from beta-lactam to macrolides (strategy S4) strongly depended on the resistance levels to both antibiotics at the country level, with this strategy being most successful when proportion of macrolide resistant (MR) was lower than the penicillin one (PNSP). This supports the concept of antibiotic cycling or switching, which have proven to be effective when adapted to account for resistance data ^29^.

Here, fitness costs values associated with specific MIC levels were calibrated to match the distribution of resistance levels in France in 2021 (2022 report of the National Pneumococci Reference Centre ^30^). The estimated fitness values ranged from 0.92 to 0.99. To our knowledge, this is the first model to incorporate a landscape of fitness costs on different levels of resistance. The resulting weighted average of the estimated fitness costs when considering the MIC distribution observed in France was 0.944. it aligns with previous estimates of the fitness cost associated with non-susceptible pneumococcus^31,32^. Regarding macrolides-resistant strains, the associated fitness cost was estimated at 0.995, indicating a low fitness burden. A study in Malawi reported the persistence of macrolide-resistance for 24 months after mass drug administration^33^, suggesting that the fitness cost associated with macrolide-resistance was insufficient to eliminate it once selection pressure was removed. These findings of persistence in macrolide-resistance are consistent with our fitness estimates.

To keep the model as simple as possible, simplifications were made to the ecology and epidemiology of *S. pneumoniae*. We did not model the emergence of resistance, focusing instead on transmission and selection as the primary drivers of resistance spread in the population. Vaccine serotype differentiation and PCV vaccinations were not accounted for in the model despite they add an additional pressure selection on strains ecology in the population. However, in most European countries the last major serotype replacement and change in pneumococcal ecology occurred following the introduction of PCV-13 more than 10 years ago (in 2010 in France). With the upcoming introduction of new vaccines such as PCV-20 and V116, which target different serotypes, their impact on future serotype selection will need to be accounted for in future modelling studies. Multi-resistance was incorporated in the model and informed by reported data in France and other countries. However the model did not include mechanisms of cross-resistance phenomena, which were less relevant for beta-lactam and macrolide resistance mechanisms^34,35^. Co-carriage was not explicitly incorporated into the model either our model considering solely carriage of the predominant strain. This choice was made to ensure model simplicity assuming that the predominant colonizing strain was the most likely to be transmitted to others and also was the one playing a significant role in infection^36^,. However, to ensure stable persistence of strains, the model allows for the possibility of a shift in the dominant strain (super colonisation). A competition cost (θ) is included to reflect this dynamic.

The mechanisms driving competition and co-existence between susceptible and resistant strains are poorly understood but play a crucial role in determining the effect of antibiotic usage on the transmission dynamics of pneumococcal carriage and disease. The model promotes coexistence and competition between susceptible and resistant strains through different plausible mechanism^37^: (i) treatment diversity, by modelling two classes of antibiotics impacting strains differently; (ii) pathogen diversity, by explicitly accounting for the diversity in non-susceptibility through MIC levels and by distinguishing resistance to both macrolide and beta-lactams; (iii) strain replacement (also called superinfection) in colonized individuals who can acquire a new strain; and (iv) fitness costs associated with resistance, by hypothesising that antibiotic-resistant strains are less transmissible than susceptible ones.

To better interpret the impact of shortages, the model was calibrated assuming that pneumococcal carriage and resistance levels were at steady state using average antibiotic consumption data for 2021 in France. Two longitudinal studies in France and the UK showed seasonal variation in pneumococcal carriage prevalence and resistance levels over at the year level^38,39^. Although this seasonality may affect the monthly magnitude of carriage prevalence, it is unlikely to alter the relative ranking of antibiotic allocation strategies or cross-country differences based on average annual effects.

Assumptions regarding the modelling of antibiotic exposure can also be discussed. Antibiotic consumption classically shows monthly fluctuation^23,40^. For simplification, we assumed here a constant exposure, therefore evaluating average yearly effects instead of short-term effects. Antibiotic exposure was also assumed to be independent of pneumococcal carriage, although children carrying pneumococci might be more likely to receive antibiotics. Some studies reported a long half-life for azithromycin^41^; however, this was estimated for 15 carbon azalides and did not concerned all macrolides^42^. Long-acting effects of macrolides was not considered here. We assumed that antibiotic-induced decolonization of susceptible strains occurred only during treatment exposure. Since the model was considered stable at the initial conditions, higher exposure among carriers or accounting for long-acting antibiotic effect would have led to different estimates for fitness cost and transmissibility. However, the predicted overall impact of the shortage scenarios would likely remain unchanged.

Additionally, the model structure should be discussed. The model does not differentiate between at-risk children and others in terms of access to antibiotics. It is possible that those at higher risk of invasive infections may be prioritized during shortages. If this had been considered, the results for resistance evolution at the population level should remain unchanged, although the predicted changes in IPD would be smaller. We have also focused our modelling on children under the age of 5, as the shortage primarily impacted paediatric antibiotic formulations worldwide. Because most of the contacts occur within the same age groups^43^, excluding other age groups is unlikely to affect our results.

Finally, data used to characterise the distribution of resistance levels were derived from IPD case-reports in France, reported by the French National Pneumococcal Reference Center (CNRP). The serotype distribution found in IPDs is generally not representative of serotype prevalence in the general population, as all pneumococcal serotypes do not have the same disease potential^44^. This may have affected our fitness cost estimates of. Moreover, country-specific differences were modelled by varying antibiotic consumption frequency and initial resistance levels, but treatment duration and global pneumococcal carriage prevalence were fixed in all simulations based on French data. This assumption should not affect our results as demonstrated by our sensitivity analysis (see Supplementary Figure 3 for the PRCC analysis) which showed that initial resistance and multi-resistance levels had the greatest impact on the recommended strategy, while treatment duration had a more limited effect. In addition, while the initial prevalence significantly impacted the outcome magnitude across all four shortage strategies, this effect was proportional, therefore not affecting the general conclusions of the paper.

Strategies can be seen both as a consequence of hypothetical recommendations implemented in response to shortages and as a concrete impact of shortages in the population. For example, strategy S1 evaluating reducing treatment consumption frequency can be interpreted either as a deliberate reduction in prescriptions by physicians aware of the shortage, or as patients struggling to obtain their treatment due to stock-outs. In the latter interpretation, strategy S1 can be used to estimate the epidemiological cost of shortages, with an additional increase of 1 to 3 IPD per 1 million children per year for a 50% shortage across European countries.

In France, a 50% beta-lactam shortage over one year could lead to an increase of 12 IPD cases among 3.8 million children under 5 years of age if the frequency of beta-lactam consumption is reduced (S1), mainly due to an increase in susceptible infections. Strategies aimed at reducing treatment duration (S2) or prescribed daily doses (S3) could also lead to additional infections, with 5 and 4 additional cases, respectively. In particular, strategy S3 would lead to an increase of 13 penicillin-non-susceptible infections. Switching to macrolides during a shortage (S4) would not affect the overall IPD incidence but would alter resistance patterns, adding 7 macrolide-resistant infection cases (see more details on the impact of shortages applied to the French context in the Supplementary Description 1).

On the one hand, focusing solely on reducing resistant infections when choosing a management strategy during antibiotic shortages might allow susceptible strains to proliferate, leading to more frequent but treatable cases of IPD. On the other hand, focusing solely on overall infections might allow resistant strains to proliferate, leading to more frequent resistant and multi-resistant IPD cases. Compared to infections by susceptible bacteria, those caused by resistant strains are associated with higher mortality^45^. This highlights the trade-off between individual benefits (usage of antibiotic to limit incidence) vs. population benefits (antibiotic use increases the resistance in the population). A complex balance therefore needs to be struck between reducing the risk of resistant infections and managing the overall disease burden.

This study was achieved in the context of increasing drug shortages, but is also in line with the World Health Organization global efforts to prevent antibiotic-resistant and multidrug-resistant infections while promoting the rational use of antibiotics^46^. Indeed, by focusing on beta-lactam shortages as part of rational antibiotic use, our proposed strategies (S1-S3) provide valuable insights for international antibiotic stewardship initiatives. Furthermore, although this study specifically investigates the impact of beta-lactam shortages, the model we developed can be extended to include resistance mechanisms of other antibiotic classes. This extension would allow the investigation of how shortages of different antibiotic classes affect pneumococcal epidemiology.

While we modelled here four distinct theorical global strategies in response to shortage this is still theoretical. In practice, physicians have to take decisions and are likely to use more complex combined strategies when confronted to shortage. In this context, understanding prescriber behaviour on the one hand and providing clear guidelines on the other hand will be key for future shortage management. In the future, this model could be used to analyse the impact of the behaviours observed during shortages, thereby enhancing our understanding of their effects on pneumococcal ecology. Additionally, it could help to guide optimal strategies during future shortages, taking into account the specific pharmaco-epidemiological context at the local level.

In conclusion, our model projections underline the importance of establishing clear public health priorities before implementing strategies to address antibiotic shortages. They also highlight the variability in optimal strategies based on country-specific pharmaco-epidemiological context. These findings demonstrate the importance of considering antibiotic stewardship on a national scale and beyond.

## MATERIALS AND METHODS

### Model Description

We developed a compartmental model (**Fig. 1**) to describe pneumococcal transmission dynamics among children under five years of age under exposure to two antibiotic classes: beta-lactams and macrolides (see Supplementary Equation. 1 for mathematical equations). Children are described according to their antibiotic exposure and colonization status (Susceptible or Carrier) and may transmit the bacteria according to the transmission rate *β*. Transmission is assumed frequency-dependent, i.e. the force of infection is proportional to the proportion of carriers in the population.

Irrespective of their carriage status, children may be exposed to either beta-lactams or macrolides, the most widely used classes of antibiotics with an effect on pneumococci (accounting for 54% and 12% respectively of antibiotic use in the French pharmaceutical sector in 2020^25^). The daily effect of this antibiotic exposure depends on the daily administrated dose and on the bacterial resistance level of the carried strain to the specific administrated antibiotic. Individuals exposed to antibiotics are assumed to be unable to acquire the sensitive strain to this class of antibiotics, reflected in the parameter *a*.

The colonization status is characterised by the strain’s resistance levels to both penicillin and macrolides. Pneumococcal resistance to beta-lactams is described according to the minimum inhibitory concentration (MIC), given that successive mutations are responsible for resistance to beta-lactams^34^. The model includes eight levels of *S. pneumoniae* penicillin susceptibility (0.063, 0.125, 0.25, 0.5,1,2,4,8 mg/L) representing increasing resistance levels. In Europe, macrolide resistance mechanism is predominantly linked to an enzymatic modification of the target (MLSB resistance)^30^, allowing only two resistance states: Susceptible or Resistant. The two molecular antibiotic resistance mechanisms are considered independent; however, we allowed strains to simultaneously carry both mechanisms more frequently than expected by chance, due to co-selection pressure over the past decades. The rate of developing an invasive pneumococcal disease (ε_*IPD*_) depends solely on carriage status and is assumed independent of antibiotic resistance level.

The cost of penicillin non-susceptibility has been shown to increase with the number of resistant alleles acquired^47^. Here, each level of *S. pneumoniae* penicillin non-susceptibility is associated with a distinct fitness cost *f*_*k*_ between 0 and 1 reducing the transmissibility of strains relatively to their fully sensitive counterpart. Macrolide resistance is also associated with its own fitness cost. For multidrug-resistant strains, the fitness cost is computed as the product of the penicillin- and macrolide-resistance associated fitness costs, and the transmission rate, given by ***f^pencillin^_k_*** × ***f^Macrolides^_k_*** × β.

At all carriage levels, competition among strains is permitted, despite colonized individual can acquire new strains through replacement (also called supercolonization). This acquisition is reduced by a competition factor *θ* applied to the transmission rate. The competition parameter *θ* reflects the extent to which carriage of one strain protects against acquisition of another^48,49^.

The code for this study is available at https://github.com/aurelmau/AntibioticShortageProject.

### Calibration of transmission parameters and antibiotic-induced decolonization rates

Given antibiotic consumption frequency, a competition factor, and decolonization rates, we fitted the transmission rate β and the fitness costs *f*_*k*_ to create a pre-shortage equilibrium state. To simultaneously estimate the transmission rate β and the fitness cost parameters *f*_*k*_ for each penicillin and macrolide resistance levels, we used the L-BFGS algorithm to maximize the likelihood by comparing the predicted pneumococcal carriage prevalence and resistance distribution in carriers with the reference distribution and prevalence, after 1 year, assuming a normal distribution.The algorithm was implemented using the optim function from the *stats* R package.

The antibiotic-induced decolonization rate **δ*^Antibiotic^_k_*** depends on the daily administered dose and the bacterial resistance level of the carried strain to the specific administered antibiotic. For beta-lactams, the parameter was calibrated to reproduce the effect of amoxicillin-clavulanate from Finnish study^50^ and for macrolides, it was calibrated to reproduce the effect of erythromycin/sulfisoxazole from a French study^51^ (see Supplementary Figure 8 and 9 for the results of calibrating the model to the data). For each of the 2 studies mentioned above, a steady state prior to antibiotic exposure was assumed by fitting the transmission rate β and fitness costs *f*_*k*_. Then, after antibiotic introduction, the antibiotic-induced decolonization parameters on susceptible and resistant strains were fitted to reproduce the change in carriage and resistance data by, by minimizing the least squares error between model predictions and reported values. We found 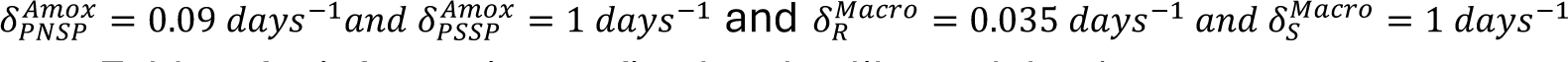 (see *PNSP PSSP R S* Supplementary Table.1 for information on fixed and calibrated data).

To better specify beta-lactam’s effects on pneumococcal strains with different levels of penicillin-non-susceptibility (based on MIC values), we assumed that the probability of decolonization decreases as MIC increases. Given that beta-lactam is time-dependent, its efficacy against pneumococcal strains was determined by the time its concentration remains above the MIC (%ft>MIC). Literature shows that for the same dosing regimens, %ft>MIC follows a sigmoid curve as a function of MIC^52^. We therefore adjusted the antibiotic-induced decolonization parameters across different resistant levels to follow a sigmoid function, ensuring that: 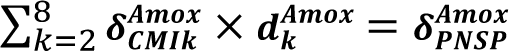 (see the adjusted values in **Table. 1**)

**Table. 1.**
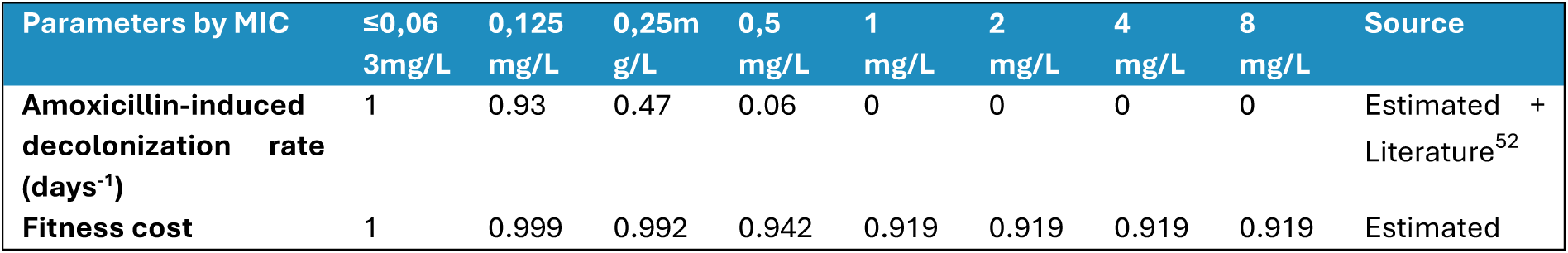
Model parameters according to penicillin minimum inhibitory concentration (MIC) values. CNRP = French reference centre for pneumococci

Using the fitted antibiotic-induced decolonization rate δ*^Antibiotic^_k_* and assuming a constant pneumococcal carriage proportion of 52%, the optimal transmission rate was estimated at β = 0.055 days^-1^.Based on the 2021 distribution from the French national reference centre for pneumococcus for otitis, the optimal fitness costs are shown in **Table. 1**.

### Other model parameters

Model parameter values are listed in Supplementary Table.2. Most values are based on data from France. The beta-lactam exposure rate is indicative of to the community beta-lactam antibiotics consumption frequency (ATC code: J01C) in children under 5 years of age in 2021. Similarly, the macrolide exposure rate is indicative of the community macrolide antibiotics consumption frequency (ATC code: J01F) in the same age group and year.

The initial *Rcr*^*init*^rate corresponds to the proportion of macrolide-resistant (*MRSP*^*init*^) strains among penicillin non-susceptible (*PNSP*^*init*^) strains. If *Rcr*^*init*^ = *MRSP*^*init*^, both resistance are independent and multi-resistance is not higher than expected by chance; if *Rcr*^*init*^ > *MRSP*^*init*^, penicillin-resistance and macrolide-resistance are not independent.

“Invasive pneumococcal diseases” (IPD) included here meningitis and bacteremia. Assuming that carriage prevalence remained constant, we calculated the daily rate of developing invasive disease using IPD incidence a French national hospital-based laboratory network (Epibac), which reported 347 cases in 2021 for children under 5 years of age^53^. This yields an estimate of ***ε_IPD_*=** 4.8 × 10^−7^ days^-1^.

### Shortage simulations

We defined four antibiotic allocation strategies under shortage, based on observed behaviours and national recommendations reported between 2022 and 2024^6,9,11,12^. Studies showed a significant drop in amoxicillin prescriptions, reaching 91% during periods of scarcity^8^. To reflect this observation, we tested different levels of shortage, corresponding to reductions in overall beta-lactam consumption frequency from 0% to 95%. Simulations were run over a 1-year period to align with the duration of shortages experienced in different countries^8^.

The four investigated strategies were: Strategy 1 (S1), reducing beta-lactam consumption frequency, which reduces the rate per child per year from 0.95 to 0.71, 0.48, 0.24, and 0.05 corresponding to 25%, 50%, 75%, and 95% shortage levels, respectively; Strategy 2 (S2), reducing beta-lactam treatment duration, which reduces the duration of exposure from 7 to 5.25, 3.5, 1.75, and 0.35 days, for the same shortage levels; Strategy 3 (S3), reducing beta-lactam daily dose, which reduces the antibiotic-induced decolonization rate among penicillin non-susceptible strains (see Supplementary Figure 10 for induced decolonization rates at different shortage levels); and Strategy 4 (S4), switching from beta-lactam to macrolide prescription, which reduces the annual beta-lactam consumption frequency per child from 0.95 to 0.71, 0.48, 0.24, and 0.05 while increasing the annual macrolide consumption frequency per child from 0.083 to 0.32, 0.56, 0.80, and 0.98, corresponding to 25%, 50%, 75%, and 95% shortage level, respectively.

We evaluated the impact of the four scenarios on seven key variables. Three related to the proportion of resistance in *S. pneumoniae* carriers: the proportion of penicillin-non-susceptible *S. pneumoniae* (PNSP), the proportion of macrolide-resistant *S. pneumoniae* (MRSP), the proportion of multidrug-resistant *S. pneumoniae* (MDRSP). Four concerning IPD outcomes : the overall invasive pneumococcal disease (IPD) incidence, the invasive pneumococcal disease susceptible (*IPD*_*SSP*_) incidence, the invasive pneumococcal disease penicillin-non-susceptible (*IPD*_*PNSP*_) incidence, the invasive pneumococcal disease macrolide-resistant (*IPD*_*MRSP*_) incidence, and the invasive pneumococcal disease multidrug-resistant (*IPD*_*MDRSP*_) incidence.

### Uncertainty analysis

Because duration of colonization is strongly variable across serotypes and age-groups, and pneumococcal prevalence strongly depends on the setting (country) or age considered, we quantified the uncertainty in model outcomes under shortage, by varying assumed values for colonization duration and carriage prevalence. Given the significant variability in these parameters across studies, we aimed to ensure that the benefits of the strategies were robust to different assumptions. In reviewed studies from the literature, duration of colonization in this age group ranged from 32 to 51 days, peaking at 43 days^54–56^, while the prevalence of carriage varied from 35% to 57%, with a peak at 52%^38,57^. Parameter values were thus drawn using a triangular distribution based on these values. Parameter combinations were then sampled using Latin Hypercube Sampling (LHS), with an LHS iteration number of 200 using the *randomLHS* function in the R package *lhs*. For each new pair of parameters, the transmissibility rate was re-calculated.

### Multivariate Sensitivity Analysis

To identify the parameters that most influence the recommended strategy in case of an beta-lactam shortage, we performed a multivariate sensitivity analysis. We computed the partial rank correlation coefficients (PRCCs) between model parameters *Rcr*^*init*^, *PNSP*^*init*^, *MRSP*^*init*^, ϕ^*Amox*^, ϕ^*Macro*^, *p*^*init*^, λ, γ^*Amox*^, and γ^*Macro*^ and four outcomes of interest after a 1-year period of 50% shortage, under the different proposed shortage management strategies. The explored outcomes were the variation and the absolute value of the variation in PNSP, MRSP, MDRSP proportions and in IPD annual incidence between shortage strategies and no shortage. Using the absolute value allowed us to focus on the amplitude in these variations, irrespective of their sign.

Parameter values were drawn according to a normal distribution with a relative standard deviation of 0.05. Parameter combinations were then sampled using LHS, with an iteration number of 500 using the *randomLHS* function in the R package *lhs*. The adjusted correlations between explored outcomes and each parameter were described using PRCCs, which were computed using the *pcc* function in the R package *sensitivity* with nboot=20^58^.

A sensitivity analysis was performed on the competition parameter θ, evaluating its impact on *S. pneumoniae* resistance under different antibiotic shortage management strategies. Five distinct values (θ=0,0.25,0.5,0.75,1) were tested to assess how varying levels of competition influence resistance dynamics. To ensure a stable pre-shortage equilibrium for each θ value, the transmission rate β and the fitness costs *f*_*k*_ were recalibrated before applying the shortages strategies.

### Simulations of specific European countries

Public health data from European countries were provided by the European Centre for Disease Prevention and control (ECDC). Specifically, data on beta-lactam and macrolide consumption frequency were obtained through the European Surveillance of Antimicrobial Consumption Network (ESAC-Net)^16^, using 2021 data. Data on pneumococcal resistance were sourced from the European Antimicrobial Resistance Surveillance Network (EARS-Net)^18^, also using 2021 data (see Supplementary Table.3 for country-specific values). EARS-Net data are based solely on invasive isolates from blood or cerebrospinal fluid. While some countries provide data from extensive national surveillance systems with high coverage, others rely on smaller subsets of local laboratories and hospitals. We included European countries with available antibiotic consumption data and at least 30 tested isolates per year.

We explored the impact of country-specific pharmaco-epidemiological conditions on our predictions. For the 20 European countries considered, we parameterised the model based on data on the proportion of penicillin non-susceptibility, macrolide resistance and penicillin-macrolide multi-resistance, as well as on beta-lactam and macrolide consumption frequency (*PNSP*^*init*^, *MR*^*init*^, *Rcr*^*init*^, Φ^*Amox*^and Φ^*Macro*^). The collected data across countries highlights strong variations in both antibiotic consumptions and resistance levels in pneumococcus (see Supplementary Figure 11 for resistance levels according to antibiotic consumption frequency in each country for ATC classes: J01C and J01F). For this analysis, other parameter values, including the contribution of children to the overall antibiotic consumption, the MIC distribution among PNSP (*d*^*Amox*^), the incidence rate of severe infections in carriers (ε_IPD_), and pneumococcal carriage prevalence (*p*^*init*^) were fixed and assumed to be equal to those sourced from French data. The calibrated fitness costs (*f*_*k*_) were assumed to be equal to those calibrated from French data, while the transmission rate (β) was re-calibrated for each country.

## Data Availability

All data produced in the present work are contained in the manuscript

