## Supplementary material for "Impact of amoxicillin shortage on pneumococcal resistance and IPD in children: evaluation of different management strategies in European countries"

**Figures**

Figure. 1 | Impact of different shortage management strategies on the evolution of pneumococcal resistance, assuming a 1-year of a 50% beta-lactam shortage, depending on the value of ${Rcr}^{init}$.

Uncertainty analysis on the assumptions regarding key parameters and initial conditions

Figure. 2 | 1-year impact of a 50% beta-lactam shortage on antibiotic resistance and the incidence of invasive pneumococcal disease (IPD), depending on the antibiotic shortage management strategy.

Figure. 3 | Multivariate sensitivity analysis of four main outcomes : partial rank correlation coefficient (PRCC) for the main model parameters under the four scenarios.

Figure. 4 | Best antibiotic allocation strategy depending on resistance level initial condition and parameters values.

Figure. 5 | Sensitivity analysis of θ competition parameter

Figure. 6 | Correlation matrix between initial conditions on country-specific pharmaco-epidemiological context and outcome variation after 1-year.

Figure. 7 | Variation in *S. pneumoniae* invasive disease incidence among children under five years of age for each resistance level to both antibiotics.

Figure. 8 | Results of calibration of the model to data on the impact of amoxicillin-clavulanate

Figure. 9 | Results of calibration of the model to data on the impact erythromycin

Figure. 10 | Amoxicillin treatment induced decolonization rate for different shortage level

Figure. 11 | Association between antibiotic consumption frequency and associated resistance across 20 European countries.

**Tables**

Table. 1 | Summary of the model parameters for the estimation of antibiotic-induced decolonization parameters

Table. 2 | Parameters of the model

Table. 3 | Country-specific pharmaco-epidemiological parameters

Table. 4 | Sensitivity analysis of theta parameter

**Descriptions**

Description. 1 | Summary of the model parameters for the estimation of antibiotic-induced decolonization parameters

**Equations**

Equation. 1 | Transmission model equations of pneumococcus colonisation

I. **Figures**

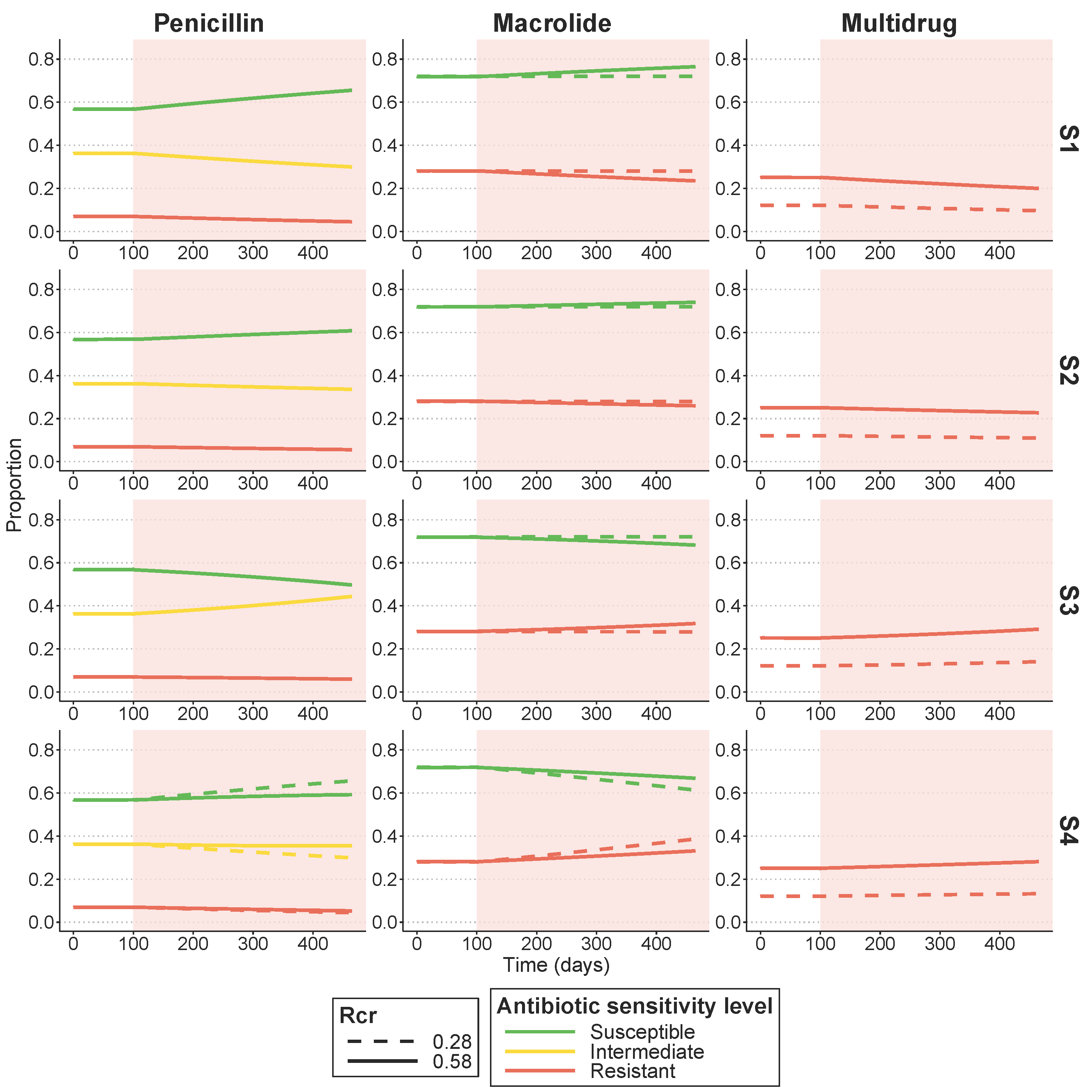

**Supplementary Figure 1. Impact of different shortage management strategies on the evolution of pneumococcal resistance, assuming a 1-year of a 50% beta-lactam shortage**, **depending on the value of** $\boldsymbol{Rcr}^{\boldsymbol{init}}$**.**

${Rcr}^{init}$ corresponds to the initial proportion of macrolide-resistant strains among penicillin non-susceptible strains (i.e., multi-resistance). If ${Rcr}^{init}={MRSP}^{init}=0.28$, both resistance are independent and multi-resistance is not higher than expected by chance; if ${Rcr}^{init}>{MRSP}^{init} (eg. {Rcr}^{init}=0.58)$, penicillin-resistance and macrolide-resistance are not independent.

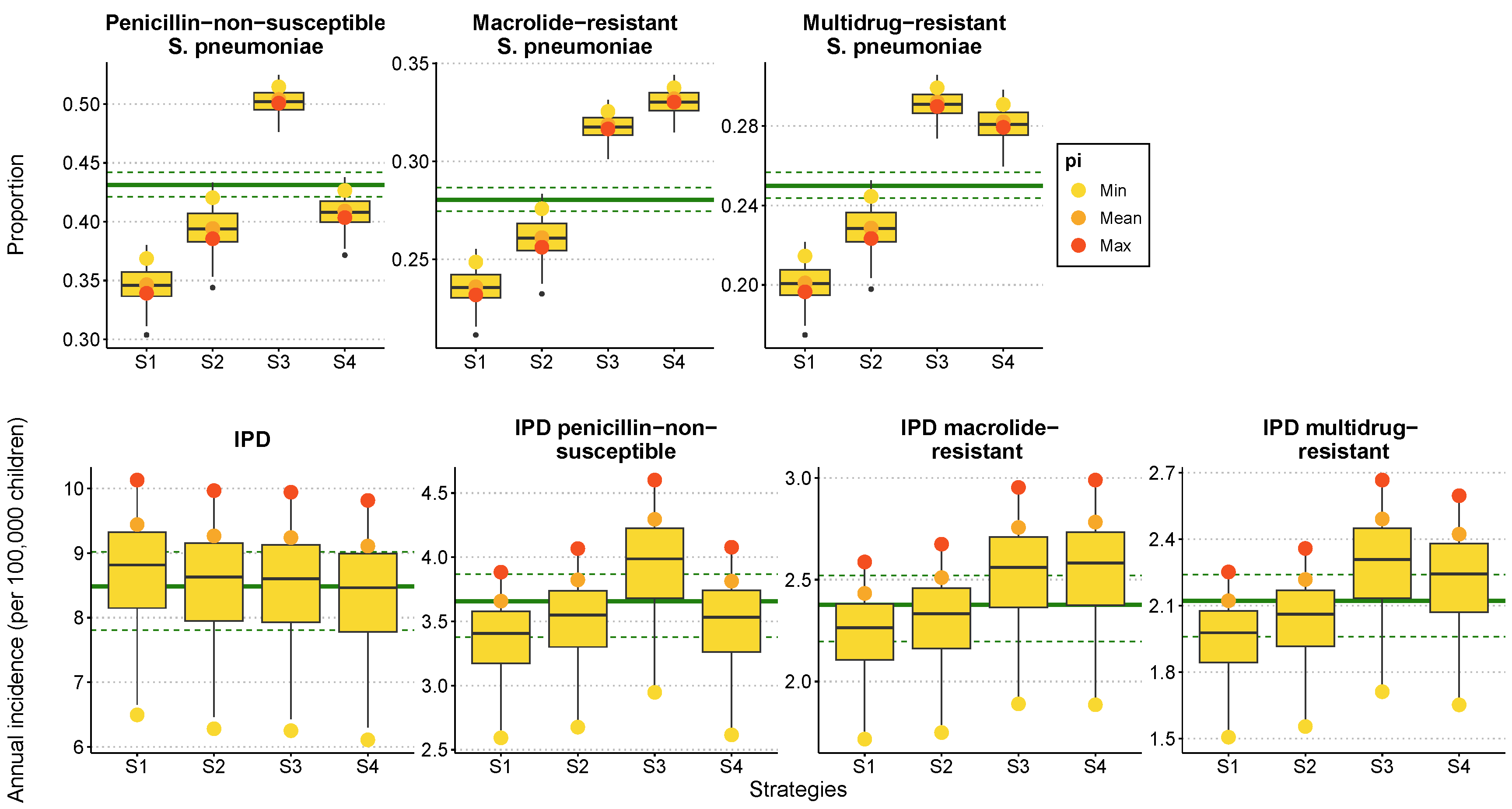

**Supplementary Figure 2. 1-year impact of a 50% beta-lactam shortage on antibiotic resistance and the incidence of invasive pneumococcal disease (IPD), depending on the antibiotic shortage management strategy.** Results are shown for three independent fixed set of parameters, the yellow, orange and red dots correspond to the outcome values for the minimum (p=0.35), average (p=0.52) and maximum (p=56) carriage prevalence values, with the carriage duration fixed at 43 days.

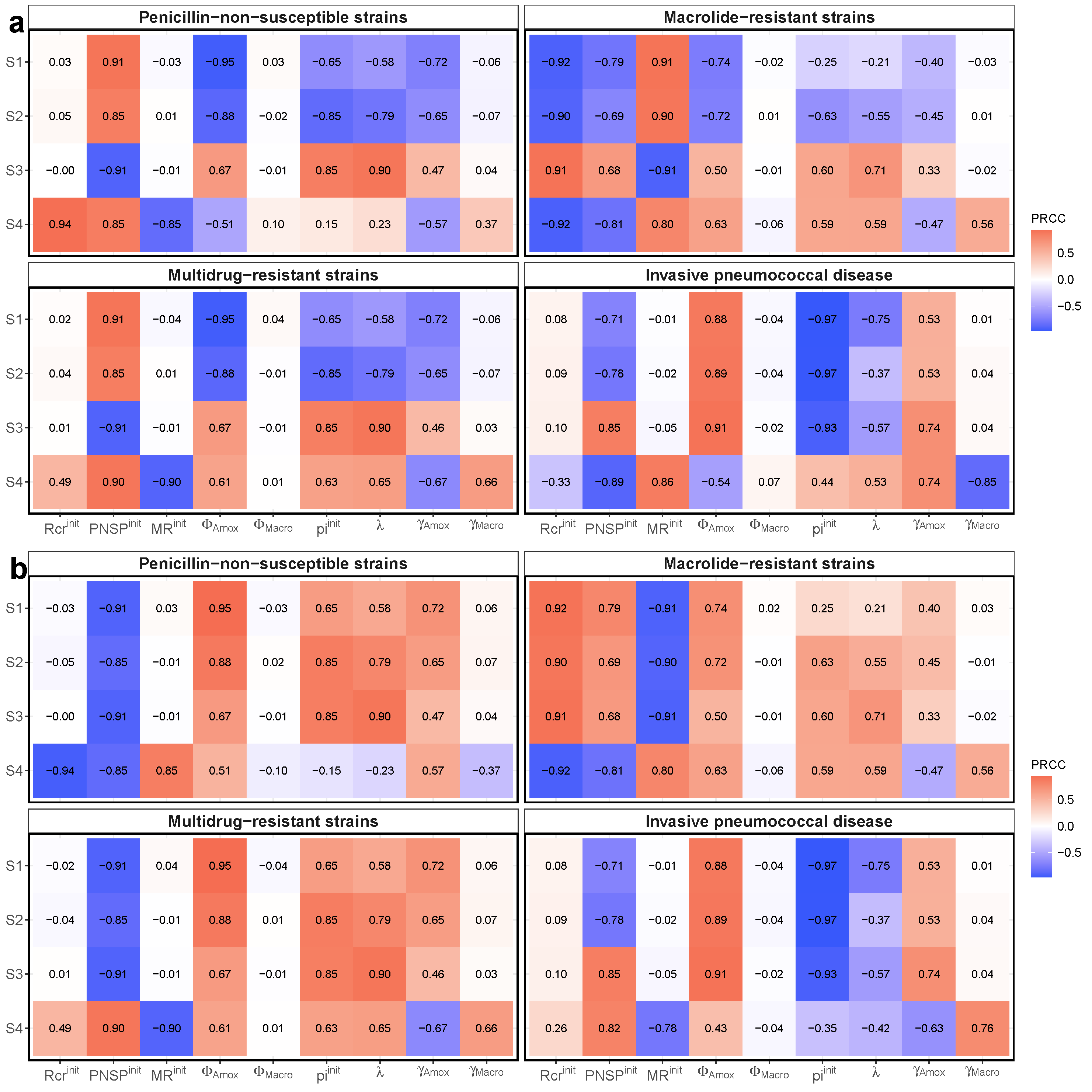

**Supplementary Figure 3. Multivariate sensitivity analysis of four main outcomes : partial rank correlation coefficient (PRCC) for the main model parameters under the four scenarios.**

**a)** PRCC of the variation of the four outcomes of interest after a 1-year period of 50% shortage, under the different proposed shortage management strategies. **b)** PRCC of the absolute value of the variation of the four outcomes of interest after a 1-year period of 50% shortage, under the different proposed shortage management strategies. A red cell indicates parameters for which, a parameter increase is associated to an increase in the variation (or absolute value of the variation)of the outcome. Blue cells indicate parameters for which a parameter increase is associated to a decrease in the variation (or absolute value of the variation). Using the absolute value allowed us to focus on the amplitude in these variations, irrespective of their sign.

In a context of shortage, allocation strategies led to a decrease or an increase of the outcome. Then, rex box indicate increases in the outcome value for positive initial value or decreases in the outcome for negative initial values. We chose to perform multivariate sensitivity analysis using the absolute value of the variations to focus on the amplitude in these variations, irrespective of their sign.

Multivariate sensitivity analysis on the absolute value of the variations show that the parameters that most impact our predictions (parameters with PRCC>0.6 or PRCC<-0.6) are ${Rcr}^{init}$ ${PNSP}^{init}$, ${MR}^{init}$, $\Phi^{Amox}$ , $p^{init}$, $\lambda$, $\gamma^{Amox}$ and $\gamma^{Macro}$. Three parameter influence patterns across explored strategies can be highlighted: (1) parameters with strong correlations across all strategies with a given outcome, such as $\Phi^{Amox}$ and $\lambda$ which consistently correlate positively with IPD variation, whereas ${pi}^{init}$ consistently correlates negatively; (2) parameters with strong correlations but varying effects across strategies for a same outcome. An example is ${PNSP}^{init}$ which is inversely correlated with IPD variation under S1 and S2, while it is positively correlated under S3 and S4; and finally (3) parameters for which the impact on a given outcome depends on the strategy. Examples are ${MR}^{init}$and $\gamma^{Macro}$ which show respectively a negative and a positive correlation with IPD when applying S4 whereas no impact in the IPD evolution could be observed with S1, S2 and S3.

This analysis highlights overall the parameters that should be considered when making the choice of optimal strategy in an beta-lactam shortage context, which are those meeting criteria (2) or (3). Specifically, key parameters are ${Rcr}^{init}$, ${MR}^{init}$, $p^{init}$ and $\lambda$ for penicillin-non-susceptible strains; ${Rcr}^{init}$, ${PNSP}^{init}$and ${MR}^{init}$ for macrolide-resistant strains; and ${PNSP}^{init}$, ${MR}^{init}$, $\gamma^{Amox}$, $\gamma^{Macro}$ for multidrug-resistant strains and severe infection incidence.

a)
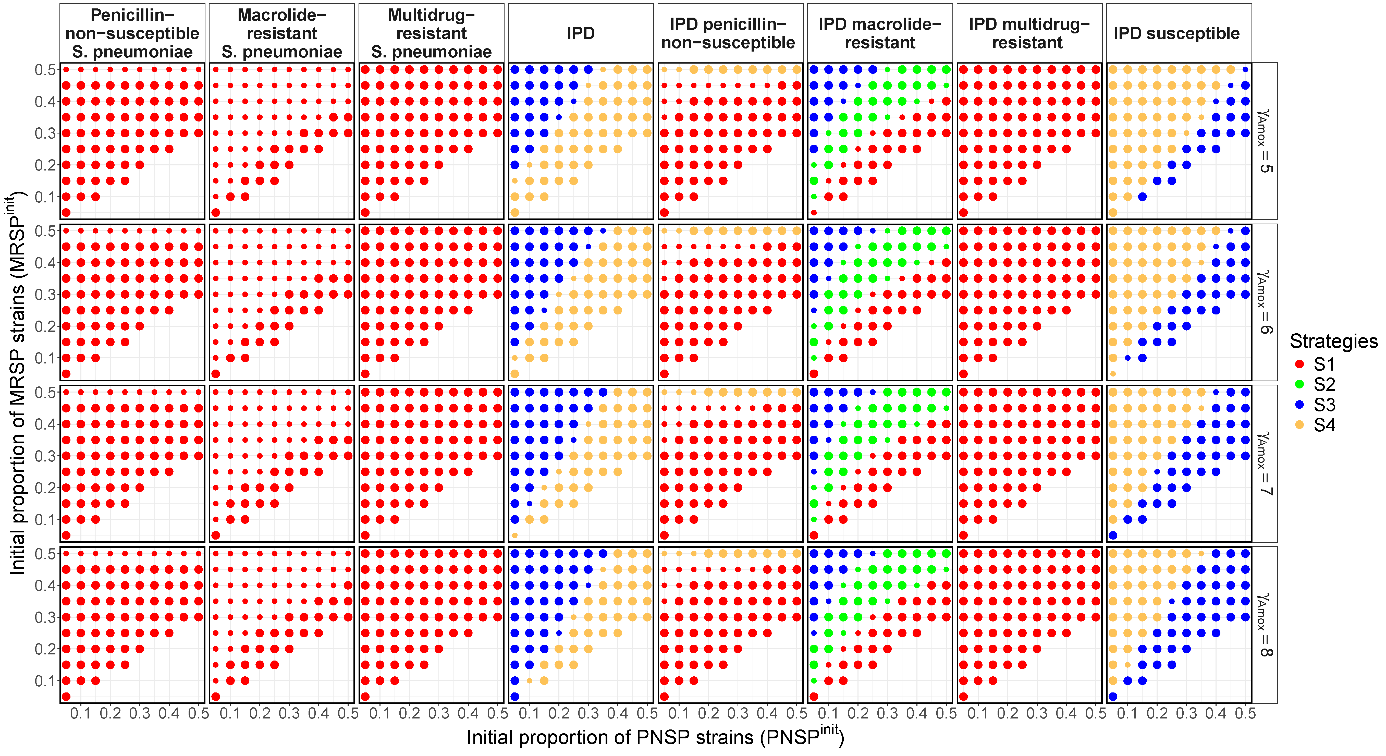

b)
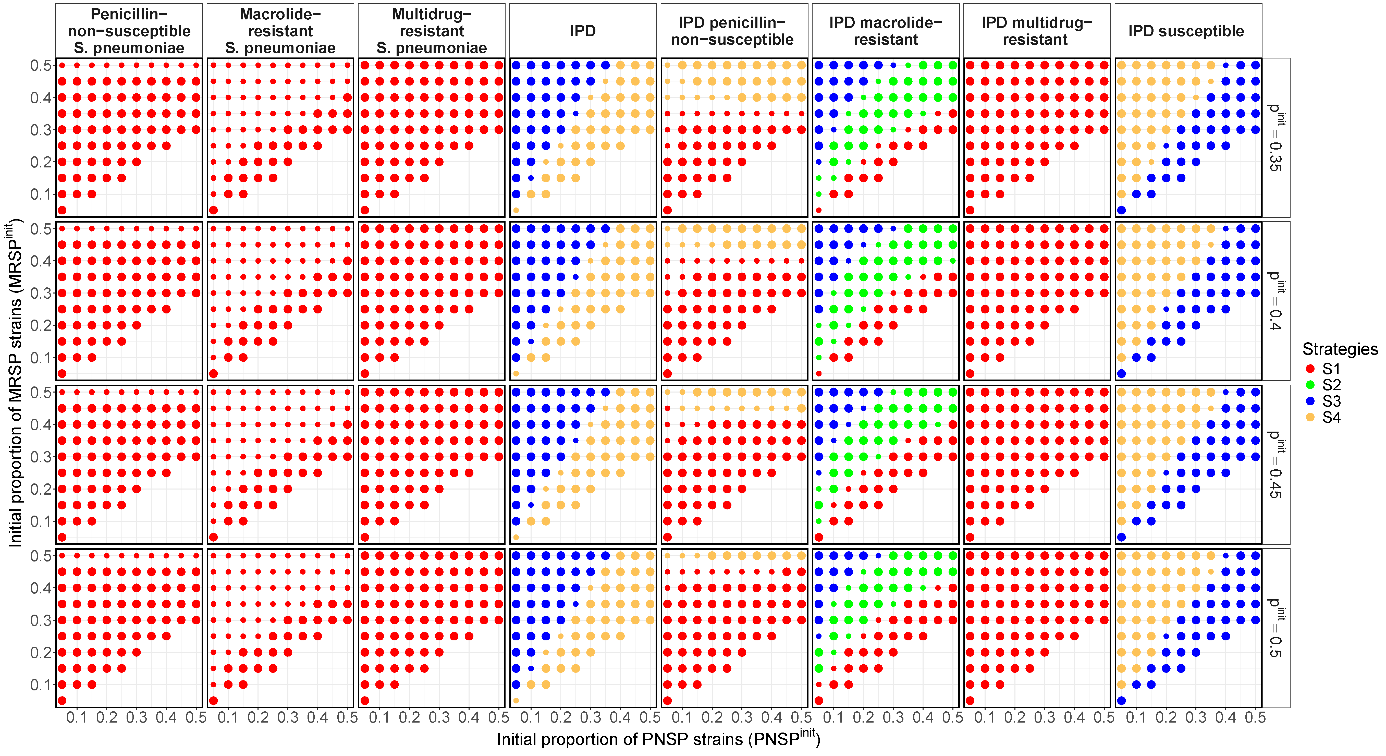

c)
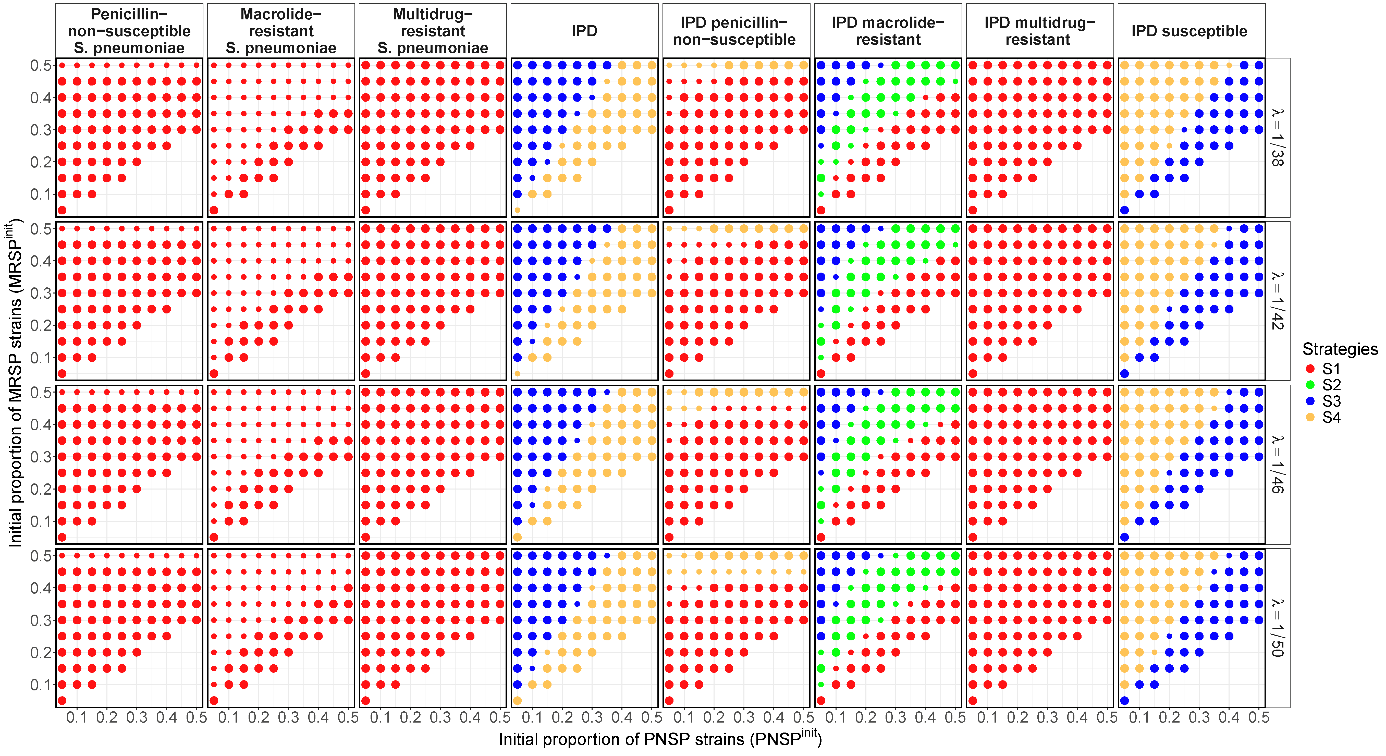

**Supplementary Figure 4. Best antibiotic allocation strategy depending on resistance level initial condition and parameters values.**

To explore the results depending in the setting, the results of the algorithm of selection of the recommended strategy for a selection of model parameters. In all figures, the selected strategy is shown for different assumptions on initial proportion of penicillin-non-susceptible *Streptococcus pneumoniae* (${PNSP}^{init}$) (x-axis) and macrolide-resistant *Streptococcus pneumoniae* (${MRSP}^{init}$) (y-axis), depending on the value of a) duration of exposure $(\gamma^{Amox}$) b) carriage prevalence ($p^{init}$) and c) duration of colonization( $1/\lambda$).

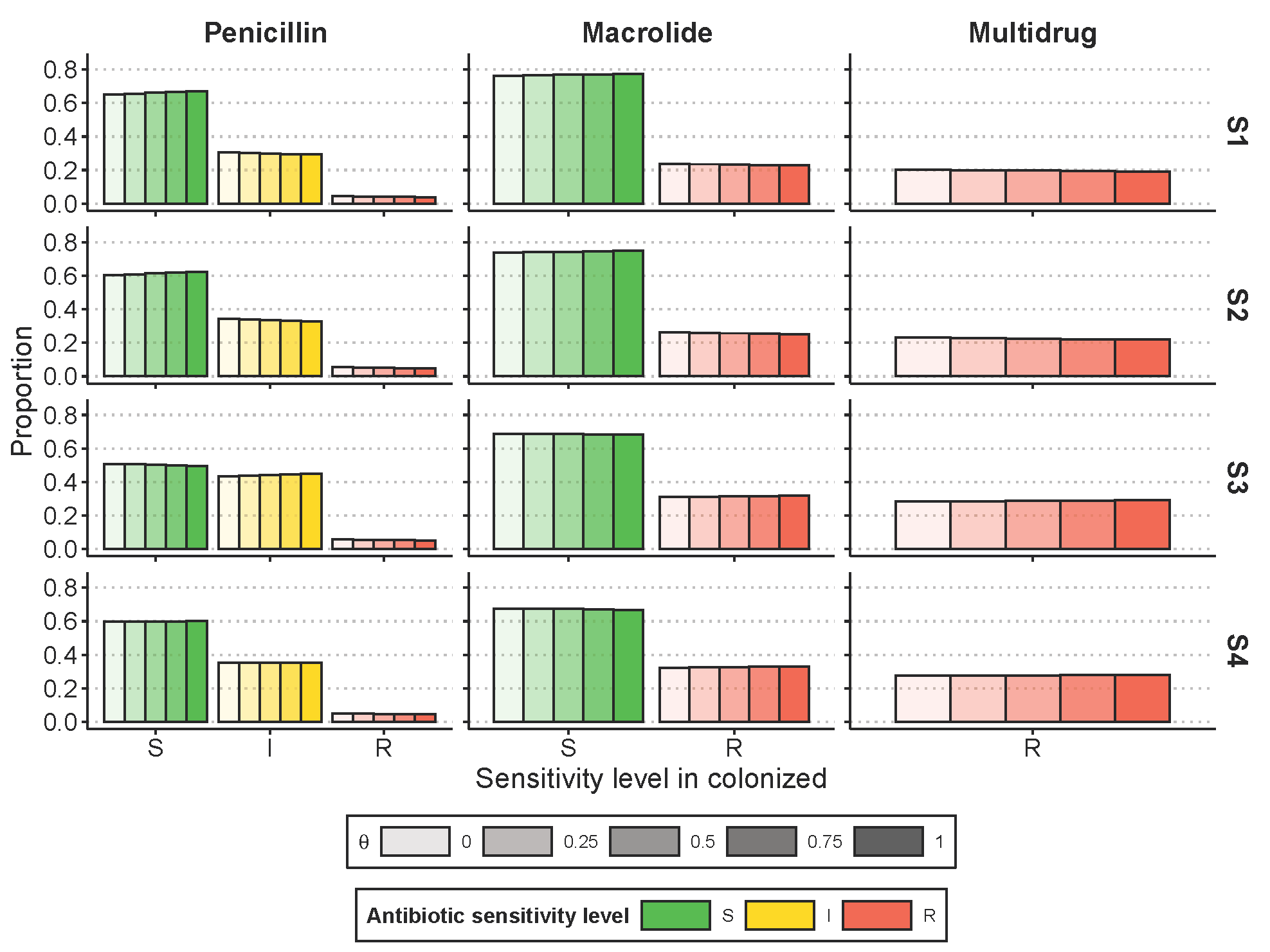

**Supplementary Figure 5. Sensitivity analysis of θ competition parameter on *S. pneumoniae* resistance, depending on the antibiotic shortage management strategy.**

We observed minimal impact from varying θ between 0 and 1 on the resistance outcome values. In fact, changes to the θ parameter primarily affected the fitted values of the fitness cost parameter. Increasing θ (which allows for more opportunities for replacement) resulted in a decrease in the fitted fitness cost, reflected by an increase in the parameter value."

| Theta/Calibrated values | $\boldsymbol{f}_{\boldsymbol{CMI}\boldsymbol{1}}^{\boldsymbol{Amox}}$ | $\boldsymbol{f}_{\boldsymbol{CMI}\boldsymbol{2}}^{\boldsymbol{Amox}}$ | $\boldsymbol{f}_{\boldsymbol{CMI}\boldsymbol{3}}^{\boldsymbol{Amox}}$ | $\boldsymbol{f}_{\boldsymbol{CMI}\boldsymbol{4}}^{\boldsymbol{Amox}}$ | $\boldsymbol{f}_{\boldsymbol{CMI}\boldsymbol{5}}^{\boldsymbol{Amox}}$ | $\boldsymbol{f}_{\boldsymbol{CMI}\boldsymbol{6}}^{\boldsymbol{Amox}}$ | $\boldsymbol{f}_{\boldsymbol{CMI}\boldsymbol{7}}^{\boldsymbol{Amox}}$ | $\boldsymbol{f}_{\boldsymbol{CMI}\boldsymbol{8}}^{\boldsymbol{Macro}}$ | $\boldsymbol{\beta}$ |
| --- | --- | --- | --- | --- | --- | --- | --- | --- | --- |
| 0 | 0.998 | 0.987 | 0.917 | 0.884 | 0.884 | 0.884 | 0.884 | 0.991 | 0.055 |
| 0.25 | 0.998 | 0.988 | 0.934 | 0.905 | 0.905 | 0.905 | 0.905 | 0.993 | 0.055 |
| 0.50 | 0.999 | 0.990 | 0.941 | 0.918 | 0.918 | 0.918 | 0.918 | 0.994 | 0.055 |
| 0.75 | 0.999 | 0.992 | 0.947 | 0.927 | 0.927 | 0.927 | 0.927 | 0.995 | 0.055 |
| 1 | 0.999 | 0.993 | 0.952 | 0.934 | 0.934 | 0.934 | 0.934 | 0.995 | 0.055 |

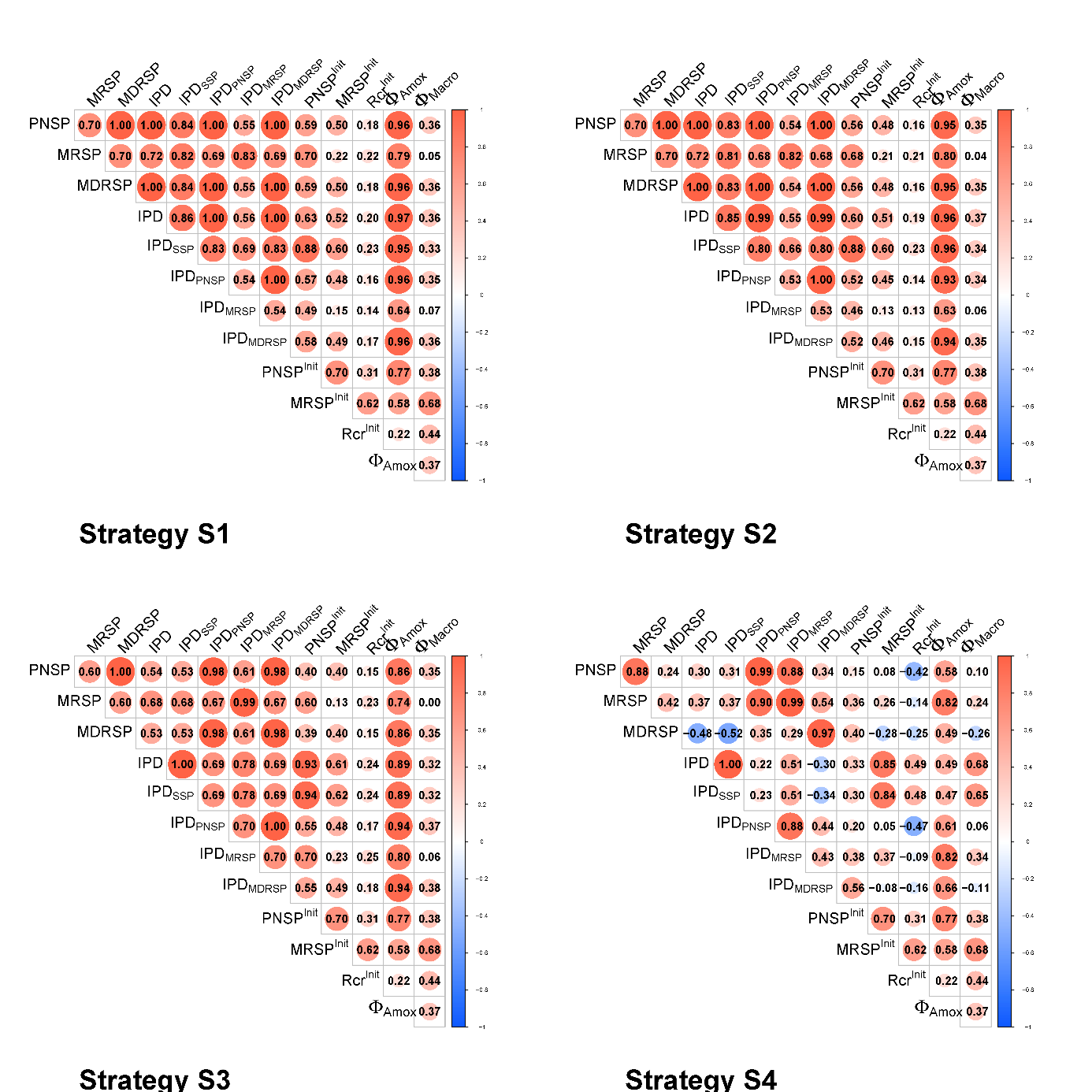

**Supplementary Figure 6. Correlation matrix between initial conditions on country-specific pharmaco-epidemiological context and outcome variation after 1-year.**

The outcome measured after 1-year of shortage are $PNSP$, $MRSP$, $MDRSP$, $IPD$, ${IPD}_{PNSP}$, ${IPD}_{MRSP}$ and ${IPD}_{MDRSP}$ absolute value of the relative variation between shortage and no-shortage. Using the absolute value allowed us to focus on the amplitude in these variations, irrespective of their sign. The parameters that varies according to the different country-specific context are ${{PNSP}^{init}, MRSP}^{init},$ ${Rcr}^{init}$, $\Phi^{Amox}$ and $\Phi^{Macro}$.

This correlation matrix is a univariate analysis that measures the strength and direction of linear relationships between two variables at a time. This analysis does not account for the influence of other variables, which can lead to misleading interpretations if confounding variables are present. This explains why the correlations found in the PRCC analysis, which is multivariate, may differ. Indeed, the PRCC determines the correlation between two variables while controlling for the influence of other variables.

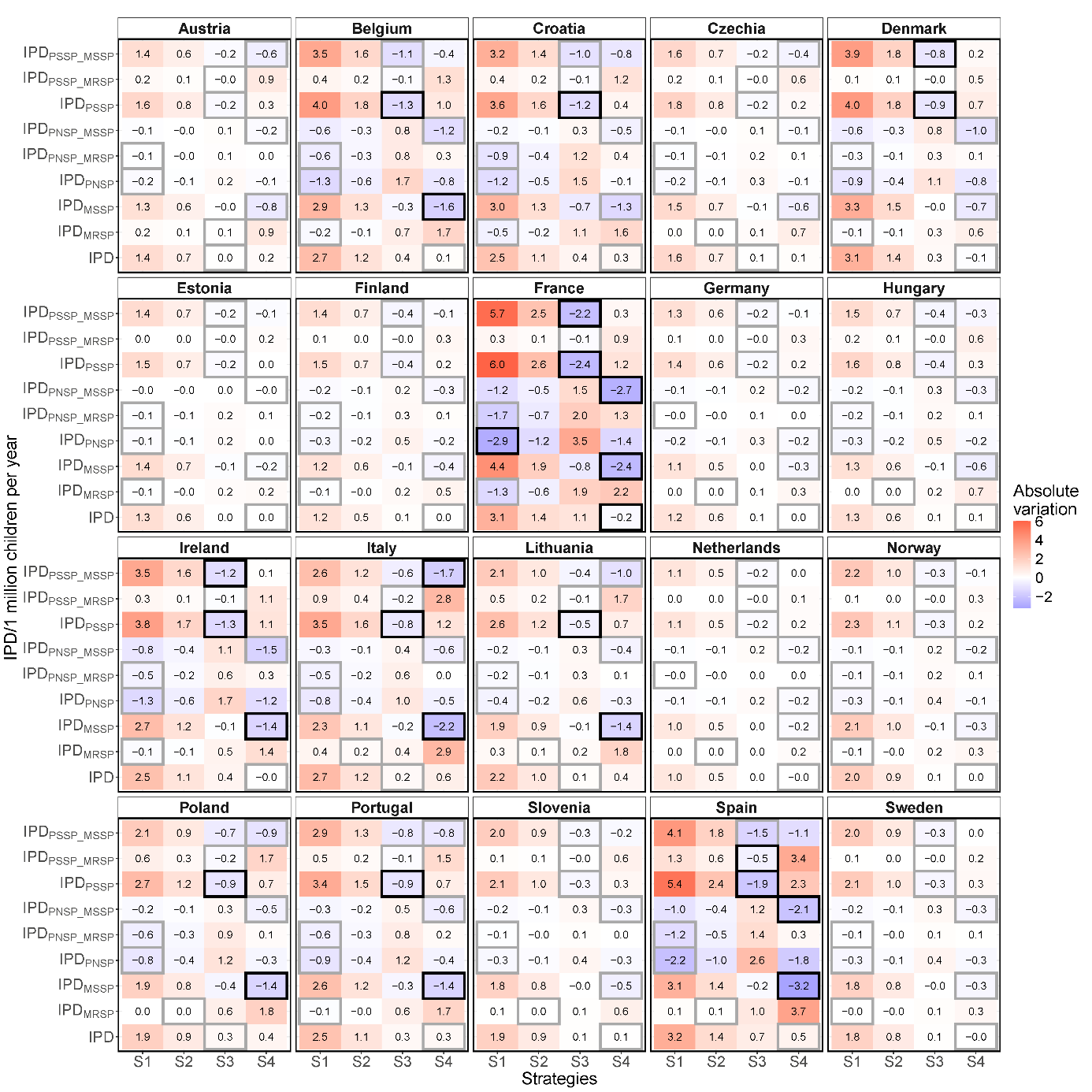

**Supplementary Figure 7. Absolute variation in *S. pneumoniae* invasive disease incidence among children under five years of age for each resistance level to both antibiotics.**

9 outcomes on the absolute variation of invasive pneumococcal disease incidence compared with no shortage are provided for each resistance levels to the two antibiotics: (a) penicillin- susceptible and macrolide-susceptible IPD incidence (${IPD}_{PSSP\_MS}$), (b) penicillin- susceptible and macrolide-resistant IPD incidence (${IPD}_{PSSP\_MR}$), (c) penicillin- susceptible IPD incidence (${IPD}_{PSSP}$) incidence, (d) penicillin-non-susceptible and macrolide-susceptible IPD incidence (${IPD}_{PNSP\_MS}$), (e) penicillin-non-susceptible and macrolide-resistant IPD incidence (${IPD}_{PNSP\_MR}$), (f) penicillin-non-susceptible IPD incidence (${IPD}_{PNSP}$) incidence, (g) macrolide-susceptible IPD incidence (${IPD}_{MS}$), (h) macrolide-resistant IPD incidence (${IPD}_{MR}$), and (i) overall IPD incidence (${IPD}_{MDR}$).

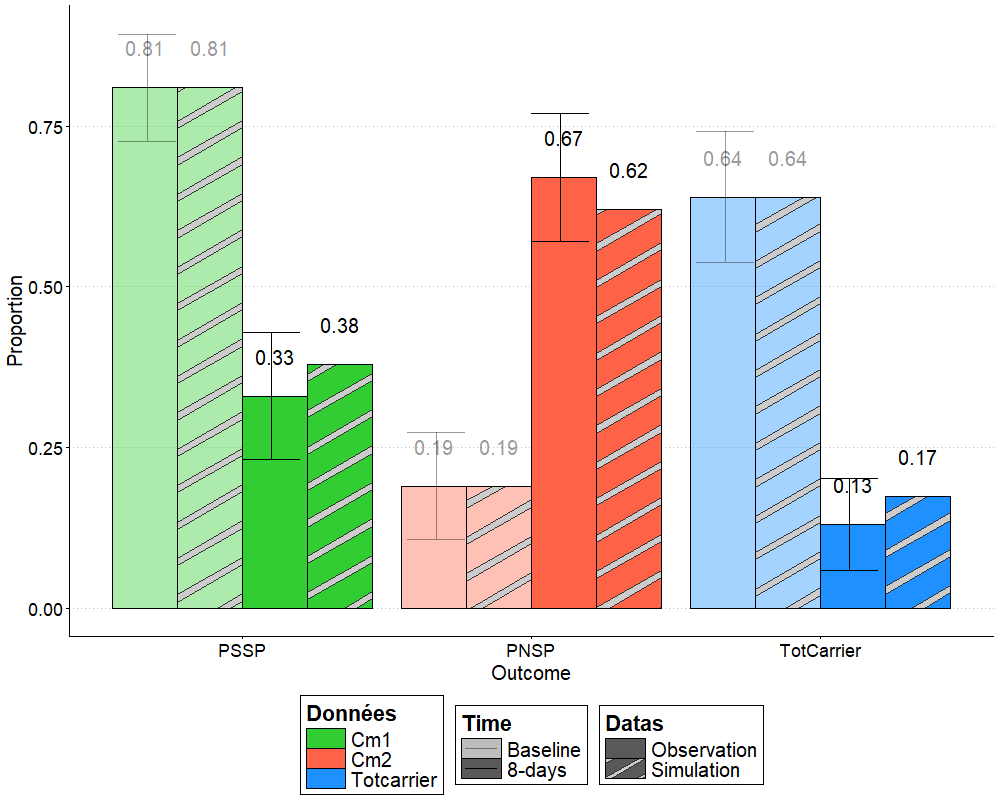

**Supplementary Figure 8. Results of calibration of the model to data on the impact of amoxicillin-clavulanate (5.7mg/kg, in three divided doses, for seven days) against *S. pneumoniae* carriage prevalence (TotCarrier) and penicillin-non-susceptible *S. pneumoniae* (PNSP) proportion, observed in 162 children aged 6-35 months, 1 day after the end of the treatment**^1^**.**

We performed these calibrations by minimizing the least squares error between model predictions of pneumococcal carriage 1 day after the end of the treatment running the Limited-memory BFGS (L-BFGS) optimization algorithm that approximates the Broyden-Fletcher-Golfarb-Shanno algorithm using box constraints^2^, limiting at 1 the antibiotic-induced decolonization rate on susceptible strains per days.

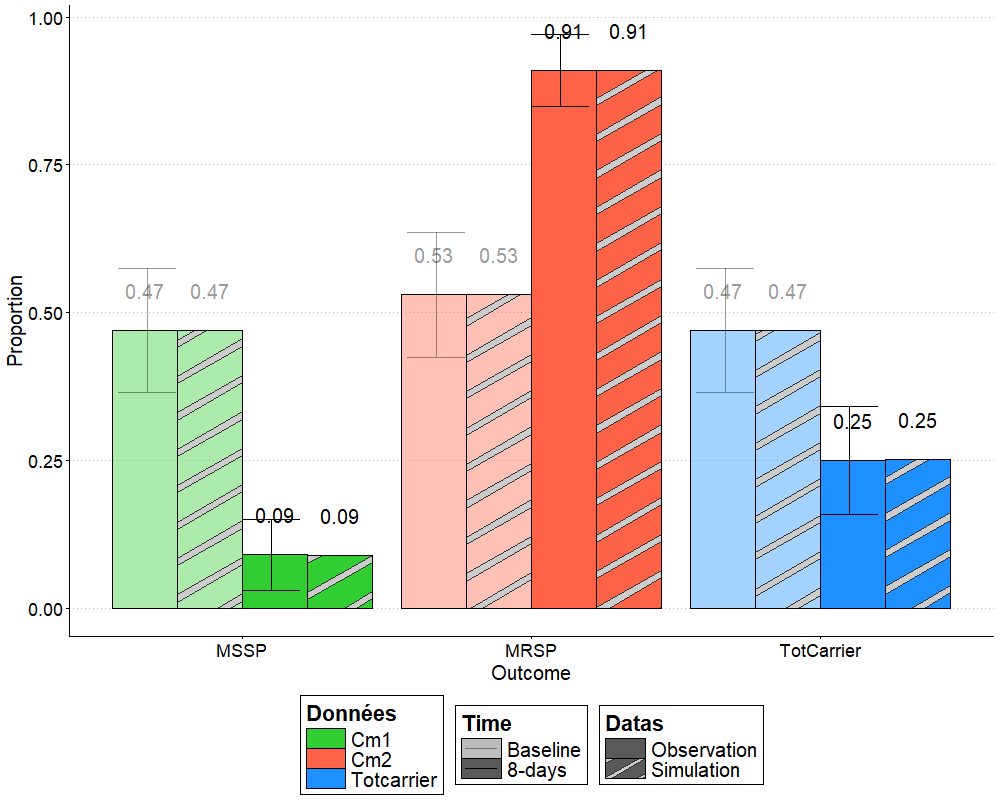

**Supplementary Figure 9. Results of calibration of the model to data on the impact erythromycin/sulfisoxazole (50mg, 150mg, in 3 divided doses, for eight days) against *S. pneumoniae* carriage prevalence (TotCarrier) and macrolide-resistant *S. pneumoniae* (MRSP) proportion, observed in 102 children aged 6-35 months, 2 days after the end of the treatment**^3^**.**

We performed these calibrations by minimizing the least squares error between model predictions of pneumococcal carriage 1 day after the end of the treatment running the Limited-memory BFGS (L-BFGS) optimization algorithm that approximates the Broyden-Fletcher-Golfarb-Shanno algorithm using box constraints^2^, limiting at 1 the antibiotic-induced decolonization rate on susceptible strains per days.

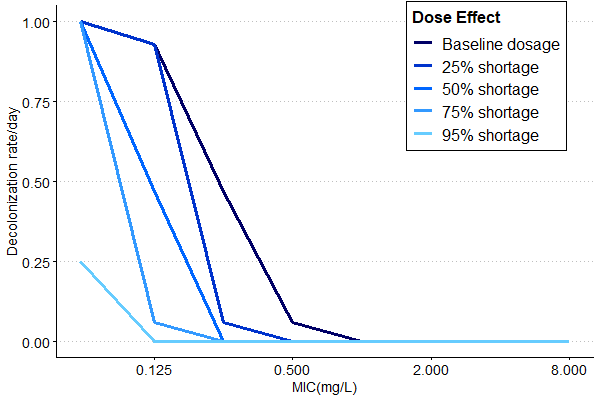

**Supplementary Figure 10. Amoxicillin treatment induced decolonization rate for different shortage level. Calibrated to follow observed change in the %ft>MIC across different MIC levels when the daily dose is reduces (de Velde, F. *et al.* Non-linear absorption pharmacokinetics of amoxicillin: consequences for dosing regimens and clinical breakpoints**^4^**).**

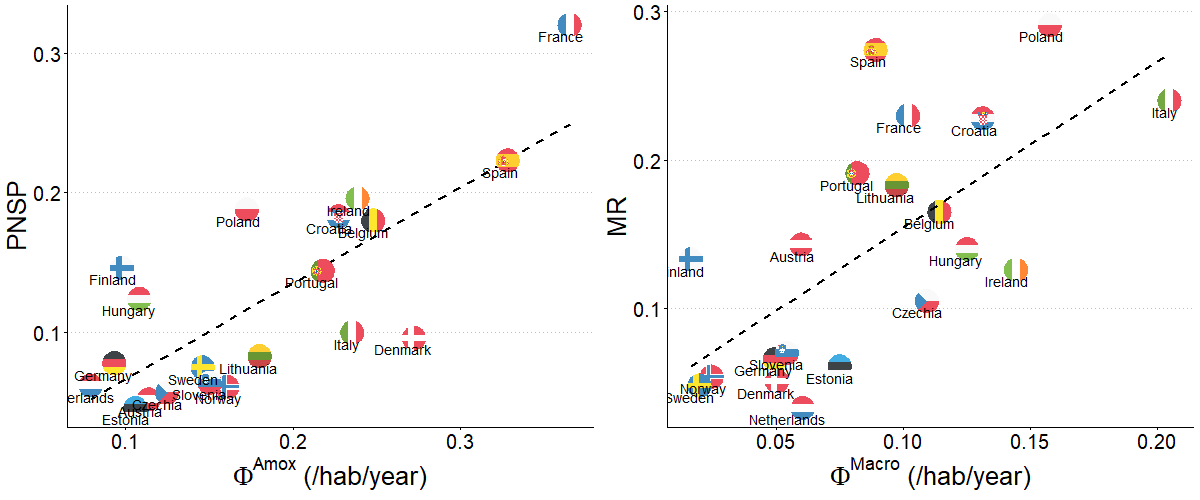

**Supplementary Figure 11. Association between antibiotic consumption frequency and associated resistance across 20 European countries.** The proportion of penicillin-non-susceptible S. pneumoniae ($PNSP$) in carriers, the proportion of macrolide-resistant S. pneumoniae ($MRSP$) in carriers, the frequency of consumption of both beta-lactams ($\Phi^{Amox}$) and macrolides ($\Phi^{Macro}$) per person per year.

Specifically, data on beta-lactam and macrolide consumption were obtained through the European Surveillance of Antimicrobial Consumption Network (ESAC-Net)^5^, using 2021 data. Data on pneumococcal resistance were sourced from the European Antimicrobial Resistance Surveillance Network (EARS-Net)^6^, also using 2021data.

II. **Tables**

|  | **Amoxicillin-clavulanate (40 and 5.7mg/kg/d, respectively, in two divided doses)** | **Erythromycin/sulfisoxazole (50 and 150mg/kg/d, respectively, in three divided doses)** | **Sources** |
| --- | --- | --- | --- |
| **Fixed data** | | |  |
| **Population size** | 146 | 86 | Amoxicillin study^1^  Erythromyci*n study* ^3^ |
| **Age of the population** | 6-35 months | 3-36 months |  |
| **Initial carriage prevalence** | 0.64 | 0.47 |  |
| **After treatment carriage prevalence** | 0.13 | 0.25 |  |
| **Initial resistance proportion** | 0.19 | 0.53 |  |
| **After treatment resistance proportion** | 0.67 | 0.91 |  |
| **Duration of treatment (**$\mathbf{1/}\boldsymbol{\gamma}$**) (days)** | 7 | 8 |  |
| **Duration of colonization (**$\boldsymbol{1/}\boldsymbol{\lambda}$**) (days)** | 43 | 43 | Literature^7,8^ |
| **Antibiotic exposure rate (**$\boldsymbol{\Phi}$**) /hab/day** | 0.007 | 0.002 | Literature^9^ |
| **Calibrated data** | | |  |
| **Rate of antibiotic-induced decolonization rate on susceptible strains** | $\delta_{PSSP}^{Amox}=1$ | $\delta_{S}^{Macro}=1$ | Calibrated |
| **Rate of antibiotic-induced decolonization rate on resistant strains** | $\delta_{PNSP}^{Amox}=0.090$ | $\delta_{R}^{Macro}=0.035$ |  |

**Supplementary Table.1 Summary of the model parameters for the estimation of antibiotic-induced decolonization parameters**

| **Parameter** | **Interprétation** | **Unit** | **Value** | **Sources** |
| --- | --- | --- | --- | --- |
| **Parameters for the <5y groups** | | | | |
| $\boldsymbol{PNSP}^{\boldsymbol{init}}$ | Initial *S. pneumoniae* penicillin-non-susceptible proportion in carriers | - | 0.43 | Pneumococcal National Reference Centre (CNRP) 2022 activity report ^10^ |
| $\boldsymbol{MR}^{\boldsymbol{init}}$ | Initial *S. pneumoniae* macrolide-resistant proportion in carriers | - | 0.28 |  |
| $\boldsymbol{Rcr}^{\boldsymbol{init}}$ | Initial proportion of macrolide-resistant strains among penicillin-non-susceptible strains | - | 0.58 |  |
| $\boldsymbol{N}$ | Population size | - | 3 850 000 | French National Institute of Statistics and Economical Studies (INSEE)^11^ |
| $\boldsymbol{\gamma}^{\boldsymbol{Amox}}$ | Rate of return to antibiotic unexposed compartment (1/duration of amoxicillin treatment | days^-1^ | 1/7 | French Health Authority (HAS)^12^ |
| $\boldsymbol{\gamma}^{\boldsymbol{Macro}}$ | Rate of return to antibiotic unexposed compartment (1/duration of macrolide treatment) | days^-1^ | 1/7 |  |
| $\boldsymbol{\Phi}^{\boldsymbol{Amox}}$ | Beta-lactam exposure rates | year^-1^ | 0.95 | Geodata in public health (Géodes)^13^ |
| $\boldsymbol{\Phi}^{\boldsymbol{Macro}}$ | Macrolide exposure rates | year^-1^ | 0.083 | Geodata in public health (Géodes)^14^ |
| $\mathbf{p}^{\mathbf{init}}$ | Carriage prevalence |  | 0.52 | Literature^15,16^ |
| $\boldsymbol{1/\lambda}$ | Duration of colonization | days | 43 | Literature^7,8^ |
| $\boldsymbol{\beta}$ | Transmissibility rate | days^-1^ | 0.056 | Estimated |
| $\boldsymbol{\varepsilon}_{\boldsymbol{IPD}}$ | Pneumococcal invasion rate | days^-1^ | $4.8\times{10}^{-7}$ | Epibac^17^ |
| $\boldsymbol{\theta}$ | Replacement penalty |  | 0.5 | Assumed |
| **Parameters by levels of resistance (k),** $\boldsymbol{if Antibiotic=Amoxicilline, k\in}\left\{ \boldsymbol{MIC}\boldsymbol{1,MIC}\boldsymbol{2,\ldots,MIC}\boldsymbol{8} \right\}\boldsymbol{if Antibiotic=Macrolide,k\in}\left\{ \boldsymbol{S,R} \right\}$ | | | | |
| $\boldsymbol{d}_{\boldsymbol{k}}^{\boldsymbol{Amox}}$ | Distribution of S.p penicillin susceptibility levels according to the MIC | - | [0.7,0.075, 0.075, 0.085, 0.09, 0.12, 0.075, 0.01] | Pneumococcal National Reference Centre (CNRP) 2022 activity report ^10^ |
| $\boldsymbol{\delta}_{\boldsymbol{k}}^{\boldsymbol{Amox}}$ | Amoxicillin-induced decolonization rate | days^-1^ | [1,0.93,0.47,0.06, 0, 0,0,0] | Literature^1,4^ and estimated |
| $\boldsymbol{f}_{\boldsymbol{k}}^{\boldsymbol{Amox}}$ | Penicillin fitness cost | - | [1, 0.999, 0.992, 0.942, 0.919, 0.919, 0.919, 0.919] | Estimated |
| $\boldsymbol{a}_{\boldsymbol{k}}^{\boldsymbol{Amox}}$ | Rate of transmission under amoxicillin exposure | - | [0, 0, 0, 1, 1, 1, 1, 1] | Assumed |
| $\boldsymbol{\delta}_{\boldsymbol{k}}^{\boldsymbol{Macro}}$ | Macrolide-induced decolonization rate | days^-1^ | [1, 0.035] | Estimated |
| $\boldsymbol{f}_{\boldsymbol{k}}^{\boldsymbol{Macro}}$ | Macrolide fitness cost | - | [1, 0.995] | Estimated |
| $\boldsymbol{a}_{\boldsymbol{k}}^{\boldsymbol{Macro}}$ | Rate of transmission under macrolide exposure | - | [0, 1] | Assumed |

**Supplementary Table.2 Parameters of the model in a French context**

| **Pays** | $\boldsymbol{PNSP}^{\boldsymbol{init}}$ | $\boldsymbol{MR}^{\boldsymbol{init}}$ | $\boldsymbol{MDR}^{\boldsymbol{init}}$ | $\boldsymbol{Rcr}^{\boldsymbol{init}}$ | $\boldsymbol{\Phi}^{\boldsymbol{Amox}}$ | $\boldsymbol{\Phi}^{\boldsymbol{Macro}}$ |
| --- | --- | --- | --- | --- | --- | --- |
| **Austria** | 0,052 | 0,143 | 0,48 | 0,02 | 0,114 | 0,060 |
| **Belgium** | 0,18 | 0,165 | 0,54 | 0,10 | 0,248 | 0,114 |
| **Croatia** | 0,183 | 0,228 | 0,86 | 0,16 | 0,227 | 0,131 |
| **Czechia** | 0,057 | 0,105 | 0,61 | 0,04 | 0,125 | 0,109 |
| **Denmark** | 0,096 | 0,051 | 0,31 | 0,03 | 0,272 | 0,050 |
| **Estonia** | 0,046 | 0,061 | 0,89 | 0,04 | 0,106 | 0,075 |
| **Finland** | 0,146 | 0,133 | 0,59 | 0,09 | 0,098 | 0,017 |
| **France** | 0,32 | 0,23 | 0,63 | 0,20 | 0,365 | 0,102 |
| **Germany** | 0,078 | 0,066 | 0,28 | 0,02 | 0,094 | 0,050 |
| **Hungary** | 0,124 | 0,14 | 0,51 | 0,06 | 0,109 | 0,125 |
| **Ireland** | 0,196 | 0,126 | 0,38 | 0,08 | 0,239 | 0,144 |
| **Italy** | 0,1 | 0,24 | 0,65 | 0,07 | 0,236 | 0,205 |
| **Lithuania** | 0,083 | 0,183 | 0,55 | 0,05 | 0,180 | 0,098 |
| **Netherlands** | 0,062 | 0,033 | 0,15 | 0,01 | 0,080 | 0,061 |
| **Norway** | 0,061 | 0,054 | 0,54 | 0,03 | 0,161 | 0,025 |
| **Poland** | 0,188 | 0,291 | 0,79 | 0,15 | 0,173 | 0,158 |
| **Portugal** | 0,144 | 0,191 | 0,68 | 0,10 | 0,218 | 0,082 |
| **Slovenia** | 0,064 | 0,07 | 0,33 | 0,02 | 0,151 | 0,054 |
| **Spain** | 0,223 | 0,274 | 0,59 | 0,13 | 0,328 | 0,089 |
| **Sweden** | 0,075 | 0,048 | 0,35 | 0,03 | 0,146 | 0,020 |

**Supplementary Table.3 Country-specific pharmaco-epidemiological parameters in the average population**

Data on pneumococcal resistance were sourced from the European Antimicrobial Resistance Surveillance Network (EARS-Net)^6^, using 2021 data. Data on beta-lactam and macrolide consumption were obtained through the European Surveillance of Antimicrobial Consumption Network (ESAC-Net)^5^, using 2021 data.

III. **Descriptions**

**Supplementary Description.1 Absolute variation in *S. pneumoniae* invasive disease incidence, considering of a 1-year of a 50% beta-lactam** **shortage** **in the French context, with 3.8 million children under 5 years of age.**

Applied to France, and considering the 3.8 million children under 5 years of age, we estimate that a 50% shortage over a year could lead, in case of reduction of beta-lactam consumption frequency (strategy S1), to an increase in overall infections by 12 [10,14] IPD cases. This includes a decrease in resistant infections of -10 [-5,-13] $\mathrm{IPD}_{\mathrm{PNSP}}$ and -5 [-2,-6] $\mathrm{IPD}_{\mathrm{MRSP}}$ cases and a larger increase in susceptible infections of 21 [17,23] $\mathrm{IPD}_{\mathrm{SSP}}$cases. Reducing beta-lactam treatment duration (strategy S2) or prescribed daily dose (strategy S3) could also increase overall infections (by 5 [5,6] and 4 [4,5] IPD cases, respectively), with S2 leading to a decrease in resistant infections (-4 [-2,-6] $\mathrm{IPD}_{\mathrm{PNSP}}$ and -2 [-1,-3] $\mathrm{IPD}_{\mathrm{MRSP}}$ cases), and S3 leading to an increase in resistant infections (13 [9,15] $\mathrm{IPD}_{\mathrm{PNSP}}$ and 7 [5,8] $\mathrm{IPD}_{\mathrm{MRSP}}$ additional cases). Finally, switching from beta-lactam to macrolide prescription in case of shortage (strategy S4) does not affect the overall IPD incidence, and results in shifting resistance patterns, with 7 [5,10] $\mathrm{IPD}_{\mathrm{MRSP}}$ and 5 [3,6] $\mathrm{IPD}_{\mathrm{MDRSP}}$additional cases but a decrease of -5 [-3,-6]$\mathrm{IPD}_{\mathrm{PNSP}}$ cases.

IV. **Equations of the model**

$$\frac{d{US}^{i}}{dt}=\mu_{0}+\lambda_{i}\sum_{l=1}^{8} \sum_{n=S}^{R} {{(UC}_{CMIl\_n}}^{i})+\gamma^{Amox}A{ES}^{i}+\gamma^{Macro}M{ES}^{i}-\left( \mu_{i}+\varphi_{i}^{Amox}+\varphi_{i}^{Macro} \right){US}^{i}-\sum_{j=1}^{4} \sum_{l=1}^{8} \sum_{n=S}^{R} {v_{i}C}_{ij}\left( \frac{f_{l}.f_{m}({{UC}_{CMIl\_n}}^{j}+{{AEC}_{CMIl\_n}}^{j}+{{MEC}_{CMIl\_n}}^{j})}{N} \right){US}^{i}$$

$$\frac{d{{UC}_{CMIk\_m}}^{i}}{dt}=\gamma^{Amox}A{{EC}_{CMIk\_m}}^{i}{+\gamma}^{Macro}M{{EC}_{CMIk\_m}}^{i}-\left( \mu_{i}+\varphi_{i}^{Amox}+\varphi_{i}^{Macro}+ \lambda_{i} \right){{UC}_{CMIk\_m}}^{i}+ \sum_{j=1}^{4} {v_{i}C}_{ij}\left( \frac{f_{k}.f_{S}\left( {{UC}_{CMIk\_m}}^{j}+{{AEC}_{CMIk\_m}}^{j}+{{AEC}_{CMIk\_m}}^{j} \right)}{N_{j}} \right){US}^{i}+\theta\sum_{j=1}^{4} {v_{i}C}_{ij}\left( \frac{f_{k}.f_{S}\left( {{UC}_{CMIk\_m}}^{j}+{{AEC}_{CMIk\_m}}^{j}+{{AEC}_{CMIk\_m}}^{j} \right)}{N_{j}} \right)\sum_{l=1}^{8} \sum_{n=S}^{R; l\_n\neq k\_m} {{UC}_{CMIl\_n}}^{i}-\theta\sum_{j=1}^{4} \sum_{l=1}^{8} \sum_{n=S}^{R; l\_n\neq k\_m} {v_{i}C}_{ij}\left( \frac{f_{l}.f_{m}({{UC}_{CMIl\_n}}^{j}+{A{EC}_{CMIl\_n}}^{j}+{M{EC}_{CMIl\_n}}^{j})}{N} \right){{UC}_{CMIk\_m}}^{i}$$

$$\frac{d{AES}^{i}}{dt}=\sum_{l=1}^{9} \sum_{n=S}^{R} (\lambda_{i}+\delta_{l}^{Amox})A{{EC}_{CMIl\_n}}^{i}+\varphi_{i}^{Amox}{US}^{i}-\left( \mu_{i}+\gamma^{Amox} \right){AES}^{i}-\sum_{j=1}^{4} \sum_{l=1}^{8} \sum_{n=S}^{R} {{a_{l}v}_{i}C}_{ij}\left( \frac{f_{l}.f_{m}({{UC}_{CMIl\_n}}^{j}+{{AEC}_{CMIl\_n}}^{j}+{{MEC}_{CMIl\_n}}^{j})}{N} \right){AES}^{i}$$

$$\frac{d{{AEC}_{CMIk\_m}}^{i}}{dt}=\varphi_{i}^{Amox}{{UC}_{CMIk\_m}}^{i}-\left( \mu_{i}+\gamma^{Amox}+ \lambda_{i}+\delta_{k}^{Amox} \right){{AEC}_{CMIk\_m}}^{i}+\sum_{j=1}^{4} {{a_{k}v}_{i}C}_{ij}\left( \frac{f_{k}.f_{S}\left( {{UC}_{CMIk\_m}}^{j}+{{AEC}_{CMIk\_m}}^{j}+{{AEC}_{CMIk\_m}}^{j} \right)}{N_{j}} \right){AES}^{i}+\theta\sum_{j=1}^{4} a_{k}{v_{i}C}_{ij}\left( \frac{f_{k}.f_{S}\left( {{UC}_{CMIk\_m}}^{j}+{{AEC}_{CMIk\_m}}^{j}+{{AEC}_{CMIk\_m}}^{j} \right)}{N_{j}} \right)\sum_{l=1}^{8} \sum_{n=S}^{R; l\_n\neq k\_m} {{AEC}_{CMIl\_n}}^{i}-\theta\sum_{j=1}^{4} \sum_{l=1}^{8} \sum_{n=S}^{R; l\_n\neq k\_m} {{a_{l}v}_{i}C}_{ij}\left( \frac{f_{l}.f_{m}({{UC}_{CMIl\_n}}^{j}+{A{EC}_{CMIl\_n}}^{j}+{M{EC}_{CMIl\_n}}^{j})}{N} \right){{AEC}_{CMIk\_m}}^{i}$$

$$\frac{d{MES}^{i}}{dt}=\sum_{l=1}^{9} \sum_{n=S}^{R} (\lambda_{i}+\delta_{n}^{Macro})M{{EC}_{CMIl\_n}}^{i}+\varphi_{i}^{Macro}{US}^{i}-\left( \mu_{i}+\gamma^{Macro} \right){MES}^{i}-\sum_{j=1}^{4} \sum_{l=1}^{8} \sum_{n=S}^{R} {{a_{m}v}_{i}C}_{ij}\left( \frac{f_{l}.f_{m}({{UC}_{CMIl\_n}}^{j}+{{AEC}_{CMIl\_n}}^{j}+{{MEC}_{CMIl\_n}}^{j})}{N} \right){MES}^{i}$$

$$\frac{d{{MEC}_{CMIk\_m}}^{i}}{dt}=\varphi_{i}^{Macro}{{UC}_{CMIk\_m}}^{i}-\left( \mu_{i}+\gamma^{Amox}+ \lambda_{i}+\delta_{m}^{Macro} \right){{MEC}_{CMIk\_m}}^{i}+\sum_{j=1}^{4} {{a_{S}v}_{i}C}_{ij}\left( \frac{f_{k}.f_{S}\left( {{UC}_{CMIk\_m}}^{j}+{{AEC}_{CMIk\_m}}^{j}+{{AEC}_{CMIk\_m}}^{j} \right)}{N_{j}} \right){MES}^{i}+\theta\sum_{j=1}^{4} a_{S}{v_{i}C}_{ij}\left( \frac{f_{k}.f_{S}\left( {{UC}_{CMIk\_m}}^{j}+{{AEC}_{CMIk\_m}}^{j}+{{AEC}_{CMIk\_m}}^{j} \right)}{N_{j}} \right)\sum_{l=1}^{8} \sum_{n=S}^{R; l\_n\neq k\_m} {{MEC}_{CMIl\_n}}^{i}-\theta\sum_{j=1}^{4} \sum_{l=1}^{8} \sum_{n=S}^{R; l\_n\neq k\_m} {{a_{m}v}_{i}C}_{ij}\left( \frac{f_{l}.f_{m}({{UC}_{CMIl\_n}}^{j}+{A{EC}_{CMIl\_n}}^{j}+{M{EC}_{CMIl\_n}}^{j})}{N} \right){{MEC}_{CMIk\_m}}^{i}$$

**Supplementary Equation.1 Transmission model equations of pneumococcus colonisation**

5.	AMC | European Centre for Disease Prevention and Control. https://qap.ecdc.europa.eu/public/extensions/AMC2_Dashboard/AMC2_Dashboard.html#eu-consumption-tab.

6.	European Centre for Disease Prevention and Control. & World Health Organization. *Antimicrobial Resistance Surveillance in Europe 2023: 2021 Data.* (Publications Office, LU, 2023).

7.	Högberg, L. *et al.* Age- and Serogroup-Related Differences in Observed Durations of Nasopharyngeal Carriage of Penicillin-Resistant Pneumococci. *Journal of Clinical Microbiology* **45**, 948–952 (2007).

8.	Ekdahl, K. *et al.* Duration of Nasopharyngeal Carriage of Penicillin-Resistant Streptococcus pneumoniae: Experiences from the South Swedish Pneumococcal Intervention Project. *Clinical Infectious Diseases* **25**, 1113–1117 (1997).

9.	Guillemot, D. *et al.* Reduction of Antibiotic Use in the Community Reduces the Rate of Colonization with Penicillin G—Nonsusceptible Streptococcus pneumoniae. *Clin Infect Dis* **41**, 930–938 (2005).

10.	Pneumococcal National Reference Centre. 2022 Activity Report. (2023).
